# Accuracy of COVID-19 self-tests with unsupervised nasal or nasal plus oropharyngeal self-sampling in symptomatic individuals in the Omicron period

**DOI:** 10.1101/2022.03.24.22272891

**Authors:** Ewoud Schuit, Roderick P Venekamp, Lotty Hooft, Irene K Veldhuijzen, Wouter van den Bijllaardt, Suzan D Pas, Vivian F Zwart, Esther B Lodder, Marloes Hellwich, Marco Koppelman, Richard Molenkamp, Constantijn Wijers, Irene H Vroom, Leonard C Smeets, Carla R S Nagel-Imming, Wanda G H Han, Susan van den Hof, Jan AJW Kluytmans, Janneke H M van de Wijgert, Karel G M Moons

## Abstract

**Background:** Performances of rapid antigen diagnostic tests (Ag-RDTs) with nasal self-sampling, and oropharyngeal plus nasal (OP-N) self-sampling, in the Omicron period are unknown.

**Methods:** Prospective diagnostic accuracy study among 6,497 symptomatic individuals aged >16 years presenting for SARS-CoV-2 testing at three test-sites. Participants were sampled for RT-PCR (reference test) and received one Ag-RDT to perform unsupervised with either nasal self-sampling (during the emergence of Omicron, and after Omicron share was >90%, phase-1) or with OP-N self-sampling (in a subsequent phase-2; Omicron share >99%). The evaluated tests were Acon Flowflex (Flowflex; phase-1 only), MP Biomedicals (MPBio), and Siemens-Healthineers Clinitest (Clinitest).

**Findings:** During phase-1, 45% of Flowflex, 29% of MPBio, and 35% of Clinitest participants were confirmatory testers (previously tested positive by a self-test at own initiative). Overall sensitivities with nasal self-sampling were 79.0% (95% CI: 74.7-82.8%) for Flowflex, 69.9% (65.1-74.4%) for MPBio, and 70.2% (65.6-74.5%) for Clinitest. Sensitivities were substantially higher in confirmatory testers (93.6%, 83.6%, and 85.7%, respectively) than in those who tested for other reasons (52.4%, 51.5%, and 49.5%, respectively). Sensitivities decreased by 6.1 (p=0.16 by Chi-square test), 7.0 (p=0.60), and 12.8 (p=0.025) percentage points, respectively, when transitioning from 29% to >95% Omicron. During phase-2, 53% of MPBio, and 44% of Clinitest participants were confirmatory testers. Overall sensitivities with OP-N self-sampling were 83.0% (78.8%-86.7%) for MPBio and 77.3% (72.9%-81.2%) for Clinitest. Comparing OP-N to nasal sampling, sensitivities were slightly higher in confirmatory testers (87.4% and 86.1%, respectively), and substantially higher in those testing for other reasons (69.3% and 59.9%, respectively).

**Interpretatio:** Sensitivities of three Ag-RDTs with nasal self-sampling decreased during Omicron emergence but was only statistically significant for Clinitest. Sensitivities were substantially influenced by the proportion of confirmatory testers. Addition of oropharyngeal to nasal self-sampling improved sensitivities of MPBio and Clinitest.

**Funding:** Dutch Ministry of Health, Welfare, and Sport.

**Research into context:** *Evidence before this study:* SARS-CoV-2 rapid antigen diagnostic tests (Ag-RDTs) require no or minimal equipment, provide a result within 15-30 minutes, and can be used in a range of settings including for self-testing at home. Self-testing may potentially lower the threshold to testing and allows individuals to obtain a test result quickly and at their own convenience, which could support the early detection of infectious cases and reduce community transmission. Real world evidence on the performance of unsupervised nasal and oropharyngeal plus nasal (OP-N) self-sampling in the Omicron variant period is needed to accurately inform end-users and policymakers. Therefore, we conducted a large prospective diagnostic accuracy study of three commercially available Ag-RDTs with self-sampling (the Acon Flowflex test, the MP Biomedicals test, and the Siemens-Healthineers Clinitest) during and after the emergence of Omicron using RT-PCR as the reference standard. Our aims were to evaluate whether the accuracies of Ag-RDTs with nasal self-sampling changed over time with the emergence of Omicron; and to determine whether addition of oropharyngeal to nasal self-sampling with the same swab yielded higher diagnostic accuracies.

*What this study adds:* The large comprehensive study was conducted in almost 6,500 participants with symptoms when presenting for routine SARS-CoV-2 testing at three public health service COVID-19 test-sites in the Netherlands. During the study, conducted between 21 December 2021 and 10 February 2022, the percentage of the Omicron variant in samples from the national SARS-CoV-2 pathogen surveillance increased from 29% in the first week to 99% in the last week of the study. The period during which the Omicron variant was dominant was divided into a nasal sampling phase (phase-1; Omicron present in >90% of surveillance samples) and an OP-N sampling phase (phase-2; Omicron share was >99%). In phase-1, 45% of Flowflex, 29% of MPBio, and 35% of Clinitest participants visited the test-site because of a positive self-test (confirmatory testers). Overall sensitivities with nasal self-sampling were 79.0% (95% CI: 74.7-82.8%) for the Flowflex, 69.9% (65.1-74.4%) for the MPBio, and 70.2% (65.6-74.5%) for the Clinitest Ag-RDT. Sensitivities were 94%, 84%, and 86%, respectively, for confirmatory testers, and 52%, 52%, and 50%, respectively, for those who had other reasons for getting tested. Sensitivities were 87.0% (79.7-92.4%), 83.1% (72.9-90.7%), and 80.0% (51.9-95.7%), respectively, in the first week, and decreased by 6.1 (p=0.16 by Chi-square test), 7.0 (p=0.60), and 12.8 (p=0.025) percentage points in the final week of the study. In Phase-2, 53% of MPBio and 44% of Clinitest participants were confirmatory testers. Overall sensitivities with OP-N self-sampling were 83.0% (78.8%-86.7%) for MPBio and 77.3% (72.9%-81.2%) for Clinitest. When comparing OP-N to nasal sampling, sensitivities were slightly higher in confirmatory testers (87.4% and 86.1%, respectively), and substantially higher in those testing for other reasons (69.3% and 59.9%).

*Implications of all the available evidence:* The sensitivities of three commercially available Ag-RDTs performed with nasal self-sampling decreased during the emergence of Omicron, but this trend was only statistically significant for Clinitest. Addition of oropharyngeal to nasal self-sampling improved the sensitivity of the MPBio and Clinitest, most notably in individuals who visited the test-site for other reasons than to confirm a positive self-test. Based on these findings, the manufacturers of MPBio and Clinitest may consider extending their instructions for use to include combined oropharyngeal and nasal sampling, and other manufacturers may consider evaluating this as well.

## Introduction

Rapid antigen diagnostic tests (Ag-RDTs) show promising performance for SARS-CoV-2 infection detection.^1-5^ Ag-RDTs require minimal equipment, provide a result within 15-30 minutes, and can be performed in a range of settings without laboratory facilities. While Ag-RDTs were initially introduced for use by trained professionals, they are currently widely available over the counter. Self-testing, without supervision of a trained professional, lowers the threshold for testing and allows individuals to obtain a test result quickly, at their own convenience. This in turn could support early detection and self-isolation of infectious cases and reduce community transmission.^6^

We previously showed that the SD Biosensor by Roche Diagnostics Ag-RDT with unsupervised nasal self-sampling had a 78.5% sensitivity in symptomatic individuals.^7^ However, since end November 2021, the Omicron variant rapidly replaced the Delta variant. Performance of Ag-RDTs for Omicron might be different due to alterations in viral proteins and infection dynamics. Initial studies comparing Omicron and Delta variants found similar sensitivities for molecular tests^8^, mixed analytical performance of lateral flow devices^9,10^, and similar real-world sensitivities for Ag-RDTs with sampling and testing by trained professionals.^11,12^ Additionally, anecdotal concerns were raised about the performance of Ag-RDTs when applying nasal self-sampling only since Omicron variant’s viral particles seem more prevalent in the throat than nose. One study indeed showed improved Ag-RDT sensitivity with combined throat and nasal sampling by trained professionals.^12^ Currently, real-world data on comparative accuracy of Ag-RDTs with unsupervised nasal or combined oropharyngeal plus nasal (OP-N) self-sampling are lacking.

Therefore, we studied the accuracy of three widely commercially available Ag-RDTs with unsupervised self-sampling during and after emergence of Omicron using RT-PCR as reference standard; to evaluate whether accuracies of Ag-RDTs with nasal self-sampling changed over time; and to quantify whether addition of oropharyngeal to nasal self-sampling yields higher diagnostic accuracies.

## Methods

The study is reported according to the STARD 2015 guidelines.^13^

### Study design and population

This large prospective diagnostic test accuracy study was embedded within the Dutch public testing infrastructure. Testing is always by RT-PCR, free-of-charge, but only available for government-approved test indications. During the study, 21 December 2021 to 10 February 2021, these were: having (i) any symptom of potential SARS-CoV-2 infection; (ii) been identified as close contact of a SARS-CoV-2 index case; (iii) tested positive on any commercially available Ag-RDT after self-sampling at own initiative (confirmatory testers); or (iv) returned from a country on the government’s list of high-risk-countries.^14^

Participants were recruited consecutively at three public health service (PHS) COVID-19 test-sites, in Rotterdam-Rijnmond (Rotterdam), Central- and Northeast Brabant (Tilburg), and West-Brabant (Roosendaal). Individuals were eligible if 16 years or older and were willing and able to sign a digital informed consent form in Dutch. Current analyses only include individuals who reported any SARS-CoV-2 infection-related symptom at the time of sampling.

The estimated share of Omicron, according to national SARS-CoV-2 pathogen surveillance, increased during the study from 29% (week 51 2021) to 99% (week 5 2022; >95% BA.1 variant).^15-17^ From 12 January 2022 onwards, the Omicron share was >90%. Most analyses, apart from the time-trend analyses, included data from the latter Omicron period. That period was further subdivided into a nasal sampling only phase (phase-1; Omicron present in >90% of surveillance samples) and an OP-N sampling phase (phase-2; Omicron share >99%).

### Inclusion procedure, specimen collection and testing

Individuals visiting one of the participating test-sites were asked by test-site staff whether they were willing to participate. If interested, they received participant information, a test-site specific Ag-RDT, and an email with a link to access study documentation. Next, trained test-site staff took a swab for routine RT-PCR testing. The RT-PCR sampling method differed slightly across test-sites; the Rotterdam and Tilburg sites used oropharyngeal and nasopharyngeal sampling, and the Roosendaal site combined oropharyngeal and nasal sampling (supplementary material 3). At all three sites, samples were tested in an off-site laboratory by RT-PCR on a Cobas 6800 or 8800 platform (Roche Diagnostics).

During the initial study weeks (in 2021 and the first one in 2022) and during phase-1 (week 2 to 3 (MPBio and Clinitest) or 5 (Flowflex) in 2022), participants received instructions to perform the Ag-RDT at home using only nasal self-sampling according to the manufacturers’ instructions. Participants received one of three Ag-RDTs: Acon Labs Flowflex COVID-19 Antigen Home Test (“Flowflex”) in Rotterdam; MP Biomedicals Rapid SARS-CoV-2 Antigen Test Card (“MPBio”) in Tilburg; and Siemens-Healthineers CLINITEST Rapid COVID-19 Antigen Test (“Clinitest”) in Roosendaal. During phase-2 (week 4 to 6 in 2022), participants in Tilburg (MPBio) and Roosendaal (Clinitest) received instructions to perform OP-N self-sampling with the same swab according to the investigator’s instructions for oropharyngeal sampling plus the manufacturer’s instructions for nasal self-sampling. We did not evaluate the Flowflex test combined with OP-N sampling because the swab provided in the Flowflex test kits was deemed not suitable for OP self-sampling. All tests are CE-marked for nasal sampling. MPBio and Clinitest were not CE-marked for OP-N sampling, but after safety checks by the quality team of the PHS West-Brabant, and consultation with in-house in-vitro diagnostic regulation experts and the Medical Ethical Committee Utrecht, both Ag-RDTs were considered safe for use with OP-N sampling.

Participants interpreted their Ag-RDT results visually according to manufacturer’s instructions, and always before they received their RT-PCR result from the PHS. Vice versa, the Ag-RDT result was not available to the laboratories that conducted the RT-PCR tests for the PHS. Participants received their RT-PCR result conform PHS routine practice to direct any further management, e.g., isolation, if applicable.

Participants were asked to complete the study procedures at home as soon as possible, and no later than within three hours of their test site visit. They were asked to first provide informed consent electronically via the participation link in the email, to subsequently perform the self-test, and to complete a short online questionnaire (supplementary material 1). Participants who did not complete this within three hours of their test-site visit were contacted by a call centre with the request to perform the self-test and complete the questionnaire as soon as possible.

Participants with a negative RT-PCR test result received an email after 10 days to complete a follow-up questionnaire (supplementary material 2) to capture any infections that were missed by the baseline RT-PCR test.

### Outcomes and statistical analyses

Primary outcomes were diagnostic accuracy (sensitivity, specificity, positive and negative predictive values with corresponding 95% confidence intervals [95% CI]) of each Ag-RDT either with nasal or with OP-N self-sampling, and RT-PCR testing as reference. Secondary outcomes were diagnostic accuracies stratified by reason for testing (confirmatory testing after a positive self-test at one’s own initiative, type of symptoms, close contact, or other reason), COVID-19 vaccination status (no vaccination or vaccinated once, twice, or thrice), having had prior SARS-CoV-2 infection, gender, and age (16-40, >40 years).

Also, we assessed whether performance of the three Ag-RDTs with nasal self-sampling changed over time during emergence of Omicron, by analysing the Ag-RDTs’ sensitivities and specificities per week. Weekly intervals were chosen because the share of the Omicron variant in The Netherlands was assessed weekly in the national pathogen surveillance.^16^ Sensitivities in the first and last week were compared by Chi-square tests.

All primary and secondary diagnostic accuracies were also determined after applying a viral load cut-off (≥5.2 log10 SARS-CoV-2 E-gene copies/mL). This was the viral load cut-off above which 95% of people with a positive RT-PCR test had a positive virus culture based on previous work.^2^ Furthermore, considering the large influence of confirmatory testers in our study populations, all analyses were repeated stratified by confirmatory testing (yes/no).

Finally, self-reported user experiences with each Ag-RDT and self-reported numbers of infections that may have been missed by baseline RT-PCR testing were assessed.

We performed complete cases analysis because the number of individuals without RT-PCR test or Ag-RDT results was very low (Figure 1).

**Figure 1.**
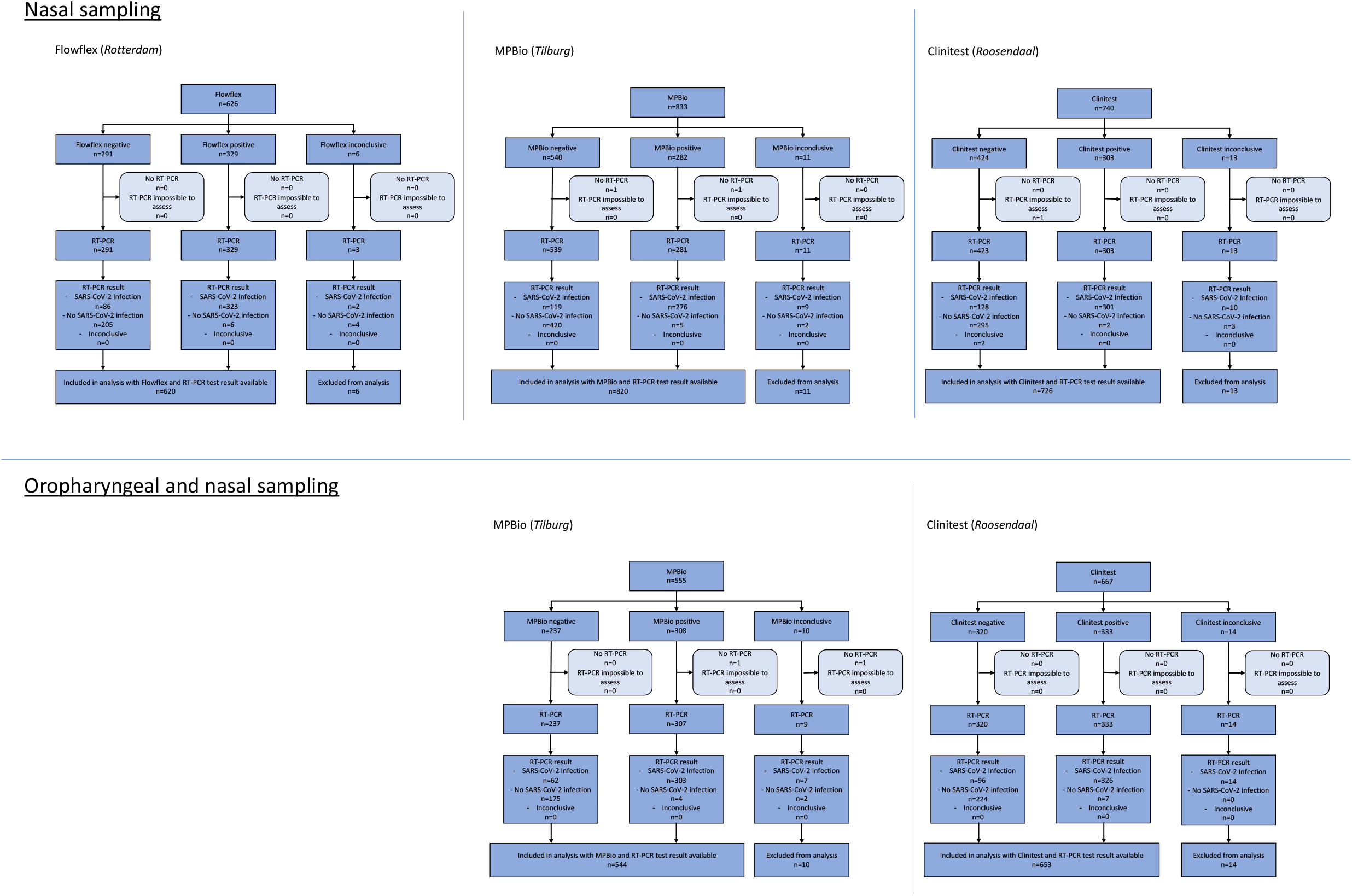
Flow of study participants in the omicron period, presented separately for nasal (top) and oropharyngeal plus nasal sampling (bottom). Flowflex = the Acon Labs Flowflex COVID-19 Antigen Home Test; MPBio = the MP Biomedicals Rapid SARS-CoV-2 Antigen Test Card; Clinitest = the Siemens-Healthineers CLINITEST Rapid COVID-19 Antigen Test. “Ag-RDT inconclusive” is a combination of Ag-RDTs that showed no control line, test tubes that were dropped, an Ag-RDTs that provided a result participants had difficulties to interpret (e.g., very light line at the “T”).

### Sample size calculation

In our previous SD-Biosensor (Roche) SARS-CoV-2 nasal Ag-RDT self-testing study, we observed a sensitivity of 79% in symptomatic participants.^7^ For the present study, we conservatively assumed sensitivities of 70% for all three Ag-RDTs irrespective of self-sampling method, with an error margin of 5%, type I error of 5% and power of 80%. Hence, we aimed for 335 positive RT-PCR tests per Ag-RDT and per self-sampling method. Since Omicron was emerging at start of the study mid-December 2021, we extended the study to ensure accrual of at least 335 positive RT-PCR tests per Ag-RDT and sampling strategy when Omicron comprised >90%.^16^

## Results

A total of 3,076 individuals participated in the Delta-Omicron transition phase prior to phase-1 (Figure S1) and a further 2,199 in phase-1 and 1,222 individuals in phase-2 (Figure 1). Participant characteristics for the Delta-Omicron transition phase are presented in Table S1 and for phases 1 and 2 in Table 1.

**Table 1.** Baseline characteristics of the symptomatic study population in the Omicron period, stratified by rapid antigen test.

| Test result available from the reference test and ... | Flowflex | MPBio |  | Clinitest |  |
| --- | --- | --- | --- | --- | --- |
| Test-site location | Rotterdam | Tilburg |  | Roosendaal |  |
| Type of sampling | Nasal | Nasal | OP-N | Nasal | OP-N |
| Inclusion dates Omicron period (>90% Omicron) | Jan 12 – Feb 3 | Jan 12 – Jan 18 | Jan 26 – Feb 10 | Jan 12 – Jan 23 | Jan 25 – Feb 8 |
| Sample size | N = 620 | N = 820 | N = 543 | N = 726 | N = 653 |
| Age [years], mean (SD) | 37 (14) | 38 (14) | 36 (13) | 39 (14) | 40 (13) |
| Range (min-max) | 16-77 | 16-85 | 16-72 | 16-81 | 16-87 |
| Sex, female, n (%)<sup>†</sup> | 357 (57.6) | 524 (63.9) | 375 (69.1) | 449 (61.8) | 437 (66.9) |
| Reported symptoms at the time of testing, n (%) | 620 (100) | 820 (100) | 543 (100) | 726 (100) | 653 (100) |
| Self-reported reasons for testing, n (%)<sup>#</sup> |  |  |  |  |  |
| Positive self-test | 279 (45.0) | 239 (29.1) | 288 (53.0) | 257 (35.4) | 290 (44.4) |
| Symptoms | 405 (65.3) | 510 (62.2) | 354 (65.2) | 459 (63.2) | 390 (59.7) |
| Close contact | 72 (11.6) | 170 (20.7) | 58 (10.7) | 144 (19.8) | 99 (15.2) |
| Other reason | 49 (7.9) | 54 (6.6) | 15 (2.8) | 29 (4.0) | 24 (3.7) |
| Vaccination status |  |  |  |  |  |
| Not vaccinated | 75 (12.1) | 68 (8.3) | 56 (10.3) | 90 (12.4) | 66 (10.1) |
| Vaccinated with at least one dose, n (%) | 545 (87.9) | 752 (91.7) | 487 (89.7) | 636 (87.6) | 586 (89.7) |
| Number of vaccinations received, n (%)<sup>§</sup> |  |  |  |  |  |
| 1 | 38 (7.0) | 86 (11.4) | 40 (8.2) | 52 (8.2) | 40 (6.8) |
| 2 | 233 (42.8) | 329 (43.8) | 184 (37.8) | 334 (52.5) | 214 (36.5) |
| 3 | 274 (50.3) | 336 (44.7) | 263 (54.0) | 248 (39.0) | 332 (56.7) |
| Unknown | 0 (0) | 1 (0.1) | 0 (0) | 2 (0.3) | 0 (0) |
| Type of initial vaccination series, n (%)<sup>§</sup> |  |  |  |  |  |
| Pfizer | 438 (80.4) | 502 (66.8) | 341 (70.0) | 410 (64.5) | 352 (60.1) |
| Moderna | 34 (6.2) | 96 (12.8) | 43 (8.8) | 116 (18.2) | 104 (17.7) |
| Astra Zeneca | 32 (5.9) | 88 (11.7) | 51 (10.5) | 62 (9.7) | 82 (14.0) |
| Janssen | 40 (7.3) | 62 (8.2) | 50 (10.3) | 45 (7.1) | 44 (7.5) |
| Unknown/Other | 1 (0.2) | 4 (0.5) | 2 (0.4) | 3 (0.5) | 4 (0.7) |
| Type of booster vaccine, n (%)<sup>§</sup> |  |  |  |  |  |
| Pfizer | 200 (36.7) | 219 (29.1) | 214 (43.9) | 168 (26.4) | 256 (43.7) |
| Moderna | 99 (18.2) | 137 (18.2) | 76 (15.6) | 93 (14.6) | 93 (15.9) |
| No booster received | 242 (44.4) | 388 (51.6) | 187 (38.4) | 366 (57.5) | 225 (38.4) |
| Unknown | 4 (0.7) | 8 (1.1) | 10 (2.1) | 9 (1.4) | 12 (2.0) |
| At least one prior SARS-CoV-2 infection, n (%) | 148 (23.9) | 185 (22.6) | 135 (24.9) | 127 (17.5) | 121 (18.5) |
| Less than 2 months ago | 14 (9.5) | 16 (8.6) | 12 (8.9) | 13 (10.2) | 11 (9.1) |
| 2 to 6 months ago | 24 (16.2) | 22 (11.9) | 25 (18.5) | 7 (5.5) | 34 (28.1) |
| 6 to 12 months ago | 62 (41.9) | 84 (45.4) | 50 (37.0) | 40 (31.5) | 33 (27.3) |
| More than 12 months ago | 48 (32.4) | 63 (34.1) | 48 (35.6) | 66 (52.0) | 43 (35.5) |
| Unknown | 0 (0) | 0 (0) | 0 (0) | 1 (0.8) | 0 (0) |
| Symptom onset, n (%) |  |  |  |  |  |
| On day of sampling | 48 (7.7) | 71 (8.7) | 59 (10.9) | 42 (5.8) | 40 (6.1) |
| A day before sampling | 239 (38.5) | 281 (34.3) | 179 (33.0) | 201 (27.7) | 178 (27.3) |
| Two days before sampling | 192 (31.0) | 257 (31.3) | 157 (28.9) | 273 (37.6) | 229 (35.1) |
| Three or more days before sampling | 140 (22.6) | 208 (25.4) | 148 (27.3) | 210 (28.9) | 205 (31.4) |
| Unknown | 1 (0.2) | 3 (0.4) | 0 (0) | 0 (0) | 1 (0.2) |
| Type of symptoms (self-reported), n (%)<sup>#</sup> |  |  |  |  |  |
| Common cold | 543 (87.6) | 717 (87.4) | 499 (91.9) | 613 (84.4) | 583 (89.3) |
| Shortness of breath | 93 (15.0) | 109 (13.3) | 87 (16.0) | 99 (13.6) | 102 (15.6) |
| Fever | 143 (23.1) | 141 (17.2) | 120 (22.1) | 162 (22.3) | 157 (24.0) |
| Coughing | 310 (50.0) | 378 (46.1) | 283 (52.1) | 386 (53.2) | 350 (53.6) |
| Loss of taste or smell | 26 (4.2) | 27 (3.3) | 20 (3.7) | 25 (3.4) | 29 (4.4) |
| Muscle ache | 137 (22.1) | 154 (18.8) | 122 (22.5) | 192 (26.4) | 152 (23.3) |
| Other symptoms | 88 (14.2) | 123 (15.0) | 85 (15.7) | 116 (16.0) | 108 (16.5) |
| Previous experience with using self-tests, n (%) | 589 (95.2) | 791 (96.7) | 535 (98.5) | 698 (96.3) | 627 (96.2) |
| Performed last self-test, n (%)<sup>§</sup> |  |  |  |  |  |
| Less than 7 days ago | 501 (85.1) | 661 (83.6) | 461 (86.2) | 588 (84.2) | 545 (86.9) |
| 1 to 4 weeks ago | 64 (10.9) | 92 (11.6) | 51 (9.5) | 75 (10.7) | 47 (7.5) |
| More than 1 months ago | 23 (3.9) | 37 (4.7) | 22 (4.1) | 35 (5.1) | 34 (5.4) |
| Unknown | 1 (0.2) | 1 (0.1) | 9 (0.2) | 0 (0) | 1 (0.2) |
| Number of ever performed self-tests, n (%) |  |  |  |  |  |
| 1-3 | 110 (18.7) | 171 (21.6) | 90 (16.8) | 193 (27.7) | 126 (20.2) |
| 4-6 | 125 (21.2) | 198 (25.1) | 128 (23.9) | 208 (29.8) | 182 (29.1) |
| 7-10 | 158 (26.8) | 181 (22.9) | 118 (22.1) | 153 (21.9) | 130 (20.8) |
| >10 | 196 (33.3) | 240 (30.4) | 199 (37.2) | 144 (20.6) | 187 (29.9) |

Flowflex=Acon Labs Flowflex COVID-19 Antigen Home Test; MPBio=MP Biomedicals Rapid SARS-CoV-2 Antigen Test Card; Clinitest=Siemens-Healthineers CLINITEST Rapid COVID-19 Antigen Test; SD=standard deviation.
<sup>#</sup> Individuals could report more than one reason for testing, or more than one symptom, respectively. Participants were asked separately whether they had symptoms on the day of study participation. All participants reported symptoms but only 60-65% reported having symptoms as a reason for testing.
<sup>§</sup> Percentage calculated as proportion of those vaccinated, or those that had previous experience in performing a self-test, respectively.

### Overall Ag-RDT accuracies with nasal self-sampling in the Omicron period

Overall sensitivities were 79.0% (74.7-82.8%) for Flowflex, 69.9% (65.1-74.4%) for MPBio, and 70.2% (65.6-74.5%) for Clinitest (Table 2, Figure 2). After applying the viral load cut-off, sensitivities increased to 85.6% (81.5-89.1%), 78.5% (73.8-82.8%), and 77.0% (72.4-81.2%), respectively (Figure S2). Specificities were >92%, positive predictive values >94%, and negative predictive values >59% for all three Ag-RDTs in all analyses (Table 2), with slightly higher specificities and PPVs for MPBio and Clinitest, and higher NPVs for Flowflex.

**Table 2.** Diagnostic accuracy parameters for the three Ag-RDTs in symptomatic individuals in the Omicron period. Values are percentages (95% confidence interval) unless stated otherwise.

|  | Sampling | N | RT-PCR test<br>positivity* [%] | Sensitivity [%]<br>(95%CI) | Specificity [%]<br>(95%CI) | PPV [%]<br>(95%CI) | NPV [%]<br>(95%CI) |
| --- | --- | --- | --- | --- | --- | --- | --- |
| **Flowflex** |  |  |  |  |  |  |  |
| Primary analysis | N | 620 | 66.0 | 79.0 (74.7-82.8) | 97.2 (93.9-98.9) | 98.2 (96.1-99.3) | 70.4 (64.8-75.6) |
| Secondary (stratified) analyses: |  |  |  |  |  |  |  |
| Viral load cut-off¶ | N | 620 | 58.2 | 85.6 (81.5-89.1) | 92.3 (88.3-95.2) | 93.9 (90.8-96.2) | 82.1 (77.2-86.4) |
| Vaccinated (at least once): |  |  |  |  |  |  |  |
| Yes | N | 545 | 64.2 | 78.6 (73.9-82.8) | 96.9 (93.4-98.9) | 97.9 (95.4-99.2) | 71.6 (65.7-77.0) |
| No | N | 75 | 78.7 | 81.4 (69.1-90.3) | 100 (79.4-100) | 100 (92.6-100) | 59.3 (38.8-77.6) |
| Previous SARS-CoV-2 infection: |  |  |  |  |  |  |  |
| Yes | N | 148 | 63.5 | 67.0 (56.6-76.4) | 98.1 (90.1-100) | 98.4 (91.6-100) | 63.1 (51.9-73.4) |
| No | N | 472 | 66.7 | 82.5 (77.9-86.6) | 96.8 (92.7-99.0) | 98.1 (95.7-99.4) | 73.4 (66.9-79.3) |
| Sex: |  |  |  |  |  |  |  |
| Female | N | 357 | 63.9 | 75.0 (68.9-80.5) | 98.4 (94.5-99.8) | 98.8 (95.9-99.9) | 69.0 (61.8-75.6) |
| Male | N | 260 | 68.5 | 83.7 (77.4-88.8) | 95.1 (88.0-98.7) | 97.4 (93.4-99.3) | 72.9 (63.4-81.0) |
| Age [years]: |  |  |  |  |  |  |  |
| 16-40 | N | 385 | 66.0 | 79.1 (73.6-84.0) | 96.2 (94.4-99.2) | 97.6 (94.4-99.2) | 70.4 (63.1-77.0) |
| >40 | N | 235 | 66.0 | 78.7 (71.4-84.9) | 98.8 (93.2-100) | 99.2 (95.6-100) | 70.5 (61.2-78.8) |
| Reason for testing was positive self-test: |  |  |  |  |  |  |  |
| Yes | N | 279 | 94.6 | 93.6 (89.9-96.2) | 80.0 (51.9-95.7) | 98.8 (96.5-99.8) | 41.4 (23.5-61.1) |
| No | N | 341 | 42.5 | 52.4 (44.0-60.8) | 98.5 (95.6-99.7) | 96.2 (89.3-99.2) | 73.7 (67.9-78.9) |
| **MPBio** |  |  |  |  |  |  |  |
| Primary analysis | N | 820 | 48.2 | 69.9 (65.1-74.4) | 98.8 (97.3-99.6) | 98.2 (95.9-99.4) | 77.9 (74.2-81.4) |
|  | OP-N | 543 | 67.2 | 83.0 (78.8-86.7) | 97.8 (94.3-99.4) | 98.7 (94.3-99.4) | 73.7 (67.6-79.2) |
| Secondary (stratified) analyses: |  |  |  |  |  |  |  |
| Viral load cut-off¶ | N | 819 | 41.5 | 78.5 (73.8-82.8) | 97.1 (95.1-98.4) | 95.0 (91.8-97.2) | 86.4 (83.2-89.2) |
|  | OP-N | 543 | 58.0 | 89.8 (86.0-92.9) | 89.5 (84.7-93.1) | 92.2 (88.6-94.9) | 86.4 (81.4-90.5) |
| Vaccinated (at least one): |  |  |  |  |  |  |  |
| Yes | N | 752 | 47.7 | 68.2 (63.2-73.0) | 99.2 (97.8-99.8) | 98.8 (96.5-99.7) | 77.4 (73.5-81.0) |
|  | OP-N | 487 | 66.9 | 82.5 (77.9-86.5) | 97.5 (93.8-99.3) | 98.5 (96.3-99.6) | 73.4 (66.9-79.2) |
| No | N | 68 | 52.9 | 86.1 (70.5-95.3) | 93.8 (79.2-99.2) | 93.9 (79.8-99.3) | 85.7 (69.7-95.2) |
|  | OP-N | 56 | 69.6 | 87.2 (72.6-95.7) | 100 (80.5-100) | 100 (89.7-100) | 77.3 (54.6-92.2) |
| Previous SARS-CoV-2 infection: |  |  |  |  |  |  |  |
| Yes | N | 185 | 35.7 | 60.6 (47.8-72.4) | 100 (96.9-100) | 100 (91.2-100) | 82.1 (74.8-87.9) |
|  | OP-N | 135 | 65.2 | 85.2 (76.1-91.9) | 97.9 (88.7-99.9) | 98.7 (92.9-100) | 78.0 (65.3-87.7) |
| No | N | 634 | 51.9 | 71.7 (66.5-76.5) | 98.4 (96.2-99.5) | 97.9 (95.2-99.3) | 76.3 (71.8-80.5) |
|  | OP-N | 408 | 67.9 | 82.3 (77.3-86.6) | 97.7 (93.5-99.5) | 98.7 (96.3-99.7) | 72.3 (65.1-78.8) |
| Sex: |  |  |  |  |  |  |  |
| Female | N | 524 | 45.0 | 70.3 (64.1-76.1) | 99.0 (97.0-99.8) | 98.2 (94.9-99.6) | 80.3 (75.8-84.3) |
|  | OP-N | 375 | 65.3 | 82.4 (77.1-87.0) | 97.7 (93.4-99.5) | 98.5 (95.8-99.7) | 74.7 (67.5-81.0) |
| Male | N | 295 | 53.9 | 69.2 (61.4-76.3) | 98.5 (94.8-99.8) | 98.2 (93.7-99.8) | 73.2 (66.2-79.5) |
|  | OP-N | 166 | 71.1 | 83.9 (76.0-80.0) | 97.9 (88.9-99.9) | 99.0 (94.6-100) | 71.2 (58.7-81.7) |
| Age [years]: |  |  |  |  |  |  |  |
| 16-40 | N | 475 | 49.5 | 75.3 (69.3-80.7) | 99.2 (97.0-99.9) | 98.9 (96.0-99.9) | 80.4 (75.4-84.8) |
|  | OP-N | 335 | 68.4 | 82.5 (77.0-87.2) | 97.2 (92.0-99.4) | 98.4 (95.5-99.7) | 72.0 (63.9-79.2) |
| >40 | N | 345 | 46.4 | 61.9 (53.9-69.4) | 98.4 (95.3-99.7) | 97.1 (91.6-99.4) | 74.9 (69.0-80.2) |
|  | OP-N | 208 | 65.4 | 83.8 (76.5-89.6) | 98.6 (92.5-100) | 99.1 (95.3-100) | 76.3 (66.4-84.5) |
| Reasons for testing was positive self-test: |  |  |  |  |  |  |  |
| Yes | N | 239 | 94.6 | 83.6 (78.1-88.2) | 84.6 (54.6-98.1) | 99.0 (96.3-99.9) | 22.9 (12.0-37.3) |
|  | OP-N | 288 | 96.2 | 87.4 (82.9-91.0) | 90.9 (58.7-99.8) | 99.6 (97.7-100) | 22.2 (11.2-37.1) |
| No | N | 581 | 29.1 | 51.5 (43.7-59.2) | 99.3 (97.9-99.8) | 96.7 (90.6-99.3) | 83.3 (79.7-86.5) |
|  | OP-N | 255 | 34.5 | 69.3 (58.6-78.7) | 98.2 (94.8-99.6) | 95.3 (86.9-99.0) | 85.9 (80.1-90.5) |
| **Clinitest** |  |  |  |  |  |  |  |
| Primary analysis | N | 726 | 59.1 | 70.2 (65.6-74.5) | 99.3 (97.6-99.9) | 99.3 (97.6-99.9) | 69.7 (65.1-74.1) |
|  | OP-N | 653 | 64.6 | 77.3 (72.9-81.2) | 97.0 (93.9-98.8) | 97.9 (95.7-99.2) | 70.0 (64.7-75.0) |
| Secondary (stratified) analyses: |  |  |  |  |  |  |  |
| Viral load cut-off¶ | N | 711 | 52.0 | 77.0 (72.4-81.2) | 97.4 (95.0-98.8) | 96.9 (94.3-98.6) | 79.6 (75.4-83.4) |
|  | OP-N | 653 | 57.3 | 83.7 (79.5-87.3) | 92.8 (89.1-95.6) | 94.0 (90.9-96.3) | 80.9 (76.2-85.1) |
| Vaccinated (at least one): |  |  |  |  |  |  |  |
| Yes | N | 635 | 58.6 | 68.5 (63.6-73.2) | 100 (98.6-100) | 100 (98.6-100) | 69.2 (64.3-73.8) |
|  | OP-N | 586 | 64.7 | 77.0 (72.5-81.2) | 98.1 (95.1-99.5) | 98.6 (96.6-99.6) | 70.0 (64.4-75.2) |
| No | N | 90 | 63.3 | 80.7 (68.1-90.0) | 93.9 (79.8-99.3) | 95.8 (85.7-99.5) | 73.8 (58.0-86.1) |
|  | OP-N | 66 | 65.2 | 79.1 (64.0-90.0) | 91.3 (72.0-98.9) | 94.4 (81.3-99.3) | 70.0 (50.6-85.3) |
| Previous SARS-CoV-2 infection: |  |  |  |  |  |  |  |
| Yes | N | 127 | 49.6 | 60.3 (47.2-72.4) | 96.9 (89.2-99.6) | 95.0 (83.1-99.4) | 71.3 (60.6-80.5) |
|  | OP-N | 121 | 49.6 | 66.7 (53.3-78.3) | 95.1 (86.3-99.0) | 93.0 (80.9-98.5) | 74.4 (63.2-83.6) |
| No | N | 599 | 61.1 | 71.9 (66.9-76.4) | 100 (98.4-100) | 100 (98.6-100) | 69.3 (64.1-74.2) |
|  | OP-N | 532 | 68.0 | 79.0 (74.4-83.1) | 97.6 (94.1-99.4) | 98.6 (96.5-99.6) | 68.6 (62.3-74.4) |
| Sex: |  |  |  |  |  |  |  |
| Female | N | 449 | 57.7 | 65.3 (59.1-71.0) | 98.9 (96.2-99.9) | 98.8 (95.8-99.9) | 67.6 (61.8-73.1) |
|  | OP-N | 437 | 60.2 | 74.9 (69.2-80.0) | 96.6 (92.6-98.7) | 97.0 (93.7-98.9) | 71.8 (65.6-77.5) |
| Male | N | 276 | 61.2 | 77.5 (70.5-83.6) | 100 (96.6-100) | 100 (97.2-100) | 73.8 (65.8-80.7) |
|  | OP-N | 213 | 73.7 | 80.9 (73.9-86.7) | 98.2 (90.4-100) | 99.2 (95.7-100) | 64.7 (53.6-74.8) |
| Age [years]: |  |  |  |  |  |  |  |
| 16-40 | N | 385 | 62.1 | 71.1 (64.9-76.8) | 99.3 (96.2-100) | 99.4 (96.8-100) | 67.8 (61.0-74.0) |
|  | OP-N | 339 | 64.9 | 79.1 (73.1-84.3) | 96.6 (91.6-99.1) | 97.8 (94.3-99.4) | 71.4 (63.8-78.3) |
| >40 | N | 341 | 55.7 | 68.9 (61.8-75.4) | 99.3 (96.4-100) | 99.2 (95.9-100) | 71.8 (65.1-77.8) |
|  | OP-N | 313 | 64.5 | 75.2 (68.7-81.0) | 97.3 (92.3-99.4) | 98.1 (94.4-99.6) | 68.4 (60.5-75.5) |
| Reason for testing was positive self-test: |  |  |  |  |  |  |  |
| Yes | N | 257 | 95.3 | 85.7 (80.7-89.8) | 91.7 (61.5-99.8) | 99.5 (97.4-100) | 23.9 (12.6-38.8) |
|  | OP-N | 290 | 96.6 | 86.1 (81.5-89.9) | 80.0 (44.4-97.5) | 99.2 (97.1-99.9) | 17.0 (7.6-30.8) |
| No | N | 469 | 39.2 | 49.5 (42.0-56.9) | 99.6 (98.1-100) | 98.9 (94.1-100) | 75.3 (70.7-79.6) |
|  | OP-N | 363 | 39.1 | 59.9 (51.3-68.0) | 97.7 (94.8-99.3) | 94.4 (87.5-98.2) | 79.1 (73.8-83.8) |

Flowflex=Acon Labs Flowflex COVID-19 Antigen Home Test; MPBio=MP Biomedicals Rapid SARS-CoV-2 Antigen Test Card; Clinitest=Siemens-Healthineers CLINITEST Rapid COVID-19 Antigen Test; NPV=negative predictive value; PPV=positive predictive value; N = nasal sampling; OP-N = combined oropharyngeal and nasal sampling. \*True SARS-CoV-2 infections based on RT-PCR test result. ¶ Defined as viral load above which 95% of individuals with a positive RT-PCR test result had a positive viral culture<sup>6</sup>, which was 5.2 log<sub>10</sub> SARS-CoV-2 E-gene copies/mL.

**Figure 2.**
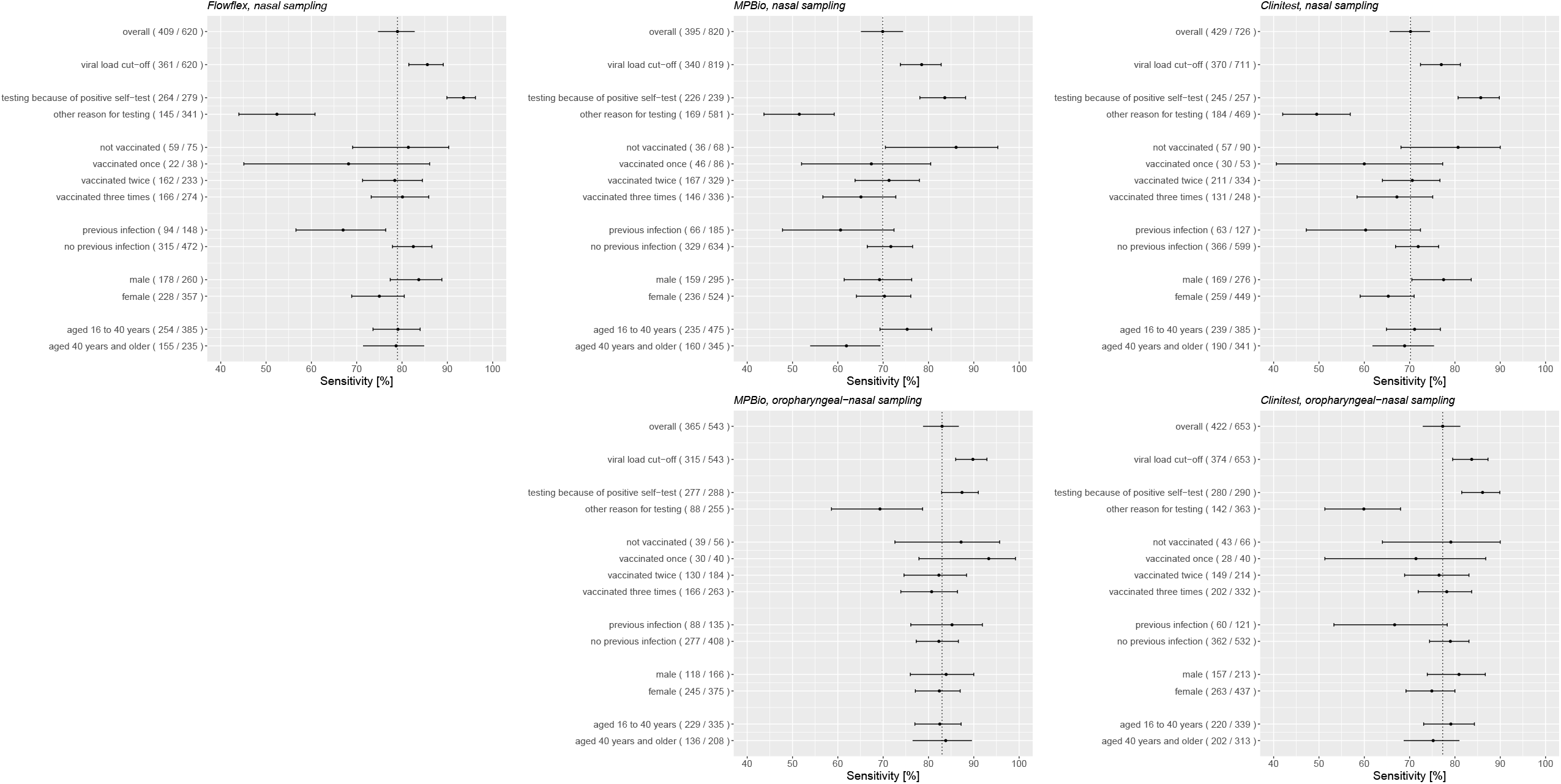
Sensitivities with 95% confidence intervals of the antigen rapid test-RT-PCR reference standard test comparisons stratified according to COVID-19 vaccination status, previous infection status, sex, and age, with nasal sampling for all three tests (top) and oropharyngeal plus nasal sampling for MPBio and Clinitest (bottom). The vertical line indicates the sensitivity of the Ag-RDT in the respective overall study population, and the number of positive RT-PCR tests out of the total or subgroup between parentheses.

Supplementary Tables S2-S4 show all 2×2 tables.

### Overall Ag-RDT accuracies with combined OP-N self-sampling in the Omicron period

Overall sensitivities were 83.0% (78.8-86.7%) for MPBio and 77.3% (72.9-81.2%) for Clinitest (Table 2, Figure 2). After applying the viral load cut-off, sensitivities increased to 89.8% (86.0-92.9%) and 83.7% (79.5-87.3%), respectively (Figure S2). Specificities, positive predictive values, and negative predictive values were for both Ag-RDTs >93%, >96%, and >75%, respectively, in all analyses (Table 2).

Supplementary Tables S5-S6 show all 2×2 tables.

### Stratified Ag-RDT accuracies with either nasal or combined OP-N self-sampling

With some differences across the three Ag-RDTs, we found lower sensitivities in participants who had had a previous SARS-CoV-2 infection, in women, and in those older than 40 (Table 2, Figure 2). Application of the viral load cut-off resulted in higher sensitivities, but all stratification trends remained similar, except that sensitivity differences by previous SARS-CoV-2 infection status were no longer significant for Flowflex (Figure S2). The largest differences in RT-PCR positivity percentages and Ag-RDTs performances were between confirmatory testers and individuals who visited the test-site for other reasons (Table 2, Figure 2). In confirmatory testers, sensitivities were 93.6% (89.9-96.2%) for Flowflex, 83.6% (78.1-88.2%) for MPBio, and 85.7% (80.7-89.8%) for Clinitest with nasal self-sampling only, and 87.4% (82.9-91.0%) for MPBio and 86.1% (81.5-89.9%) for Clinitest with combined OP-N self-sampling (Table 2, Figure 2). In individuals who tested for other reasons, sensitivities were 52.4% (44-60.8%) for Flowflex, 51.5% (43.7-59.2%) for MPBio, and 49.5% (42.0-56.9%) for Clinitest with nasal self-sampling only, and 69.3% (58.6-78.7%) for MPBio and 59.9% (51.3-68.0%) for Clinitest with OP-N self-sampling. Diagnostic accuracies stratified by all reasons for testing are presented in Table S7.

Repeating all primary and secondary analyses in confirmatory and non-confirmatory testing participants separately indicated no distinctly different trends in sensitivities across subgroups (Figure S3 for nasal sampling and Figure S4 for OP-N sampling). Differences across subgroups were less pronounced in the confirmatory testers than in non-confirmatory testers, with much higher sensitivities among confirmatory testers in all strata.

### Ag-RDT accuracy changes over time during emergence of Omicron with nasal self-sampling only

Sensitivities of all three Ag-RDTs were highest during the first week (Figure 3) when the Omicron share was 28.6%: 87.0% (79.7-92.4%) for Flowflex, 80.0% (51.9-95.7%) for MPBio, and 83.1% (72.9-90.7%) for Clinitest. With emergence of Omicron, sensitivities decreased by 6.1 percentage points for Flowflex (Chi-square test statistic 2.0; p-value=0.16), 7.0 for MPBio (Chi-square test statistic 0.28; p-value=0.60), and 12.8 for Clinitest (Chi-square test statistic 5.0; p-value=0.025). Specificities varied between 93.2% and 99.6% over time. Application of the viral load cut-off resulted in higher sensitivities but all trends over time remained similar (Figure 3).

**Figure 3.**
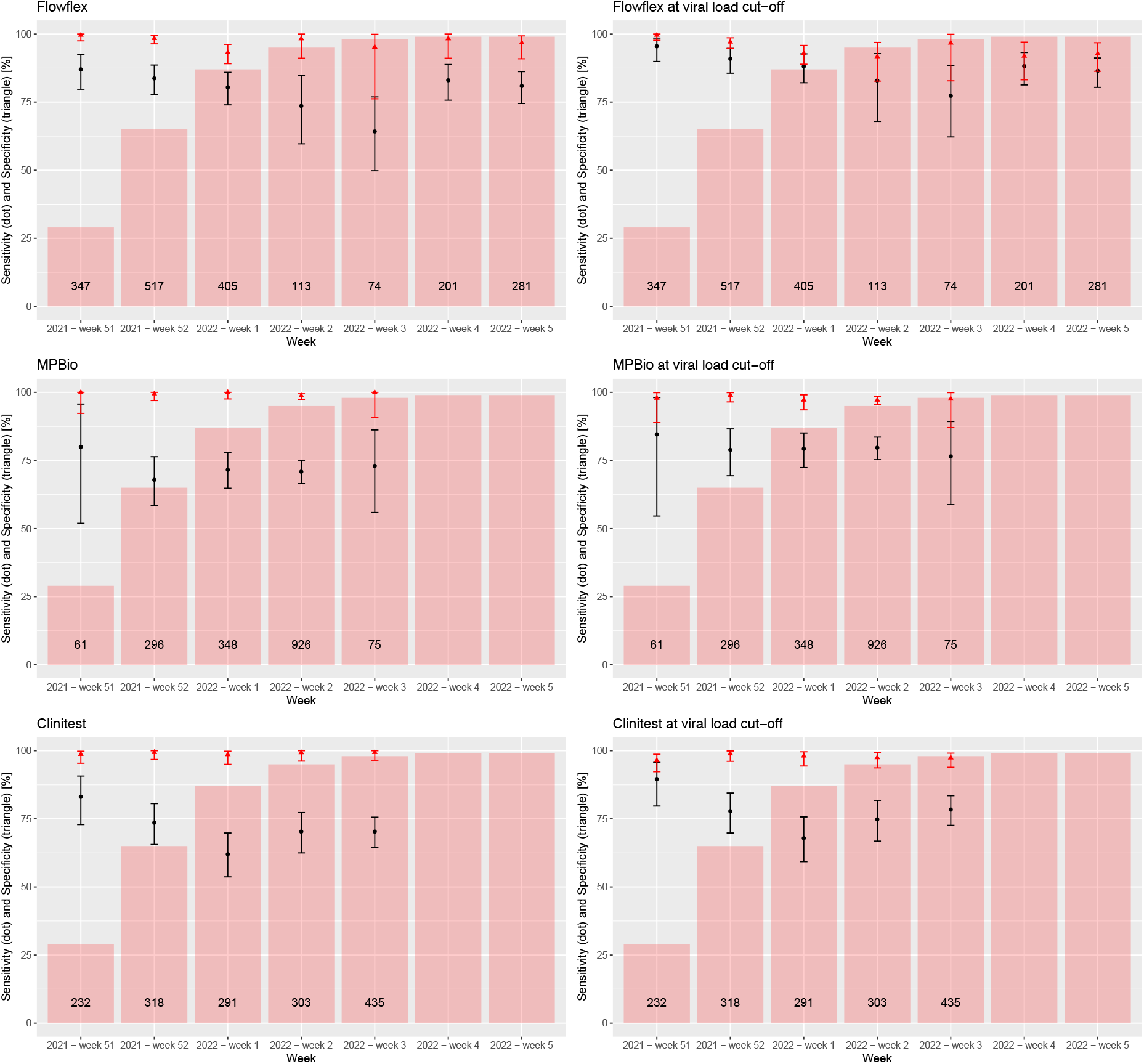
Sensitivities (dots) and specificities (triangles) with 95% confidence intervals of the antigen rapid test with nasal sampling-RT-PCR reference standard test comparison by week of inclusion, before (left) and after application of a viral load cut-off (right). The red bars indicate the percentage of the Omicron variant according to the national pathogen surveillance, while the numbers indicate the number of participants included in each week.

Stratification of the time trend analysis by reason for testing revealed that these time trends were similar but more pronounced in non-confirmatory testers than in confirmatory testers, although confidence intervals are wide (Figure S5).

### User experiences and 10-day follow-up

Information on user experiences and positive RT-PCR tests during the 10-day follow-up period are presented in Tables S8 and S9, respectively.

## Discussion

This large diagnostic accuracy evaluation of three commercially available Ag-RDTs in symptomatic individuals showed overall sensitivities ranging from 70% to 79%, increasing to 77% to 85% when a viral load cut-off was applied, for unsupervised nasal self-sampling during a period in which Omicron was dominant. Emergence of Omicron was associated with a 6 to 13 percentage points decline in overall Ag-RDTs’ sensitivities when combined with nasal self-sampling, although this trend was only significant for Clinitest. The overall sensitivities of the MPBio and Clinitest Ag-RDTs increased from 70% to 83% and 70% to 77%, respectively, when combined with OP-N self-sampling instead of nasal self-sampling only. All above-mentioned sensitivities were substantially higher in confirmatory testers than in those who visited test-sites for other reasons.

Our pre-Omicron studies, and when less than 5% of participants were confirmatory testers, found sensitivities of 72-83% for three different Ag-RDTs when performed by trained professionals, and 78.5% for the SD Biosensor Ag-RDT when performed with unsupervised self-sampling from the nose.^7,18,19^ The sensitivities that we found in the first week of the current study, when Delta was still highly dominant, were similar (Flowflex 87%, MPBio 80%, and Clinitest 83%), although the percentage of confirmatory testers in this study was much higher (21%-24%). However, sensitivities declined to 80%, 70%, and 70%, respectively, in the Omicron period. Recent studies from the USA and Italy that evaluated Ag-RDTs when Omicron was dominant, found comparable sensitivities of 74% (128/173 RT-PCR positives) and 82% (126/154 RT-PCR positives), but sampling was performed by professionals and sample sizes were smaller.^11,12^

We postulate several potential reasons for the somewhat lower sensitivities with Omicron. First, mutations in the Omicron’s Nucleocapsid (N) protein, the target of Ag-RDTs, could influence binding efficiency of antibodies used in the Ag-RDTs. However, analytical sensitivity based on isolated Omicron and Delta virus generally appeared similar.^9^ Second, the proportion of confirmatory testers, who obviously have a higher a-priori chance of testing positive on the Ag-RDT, may have fluctuated over time and by test-site, although our time trend analyses did not confirm this hypothesis (see below). Third, a larger proportion of individuals over time have experienced a SARS-CoV-2 infection, which may have affected Ag-RDT performance (see below). Fourth, during the study period the participating test-sites and laboratories experienced increasing requests for testing, exceeding the maximum capacity of the Dutch testing infrastructure. As a result, the exposure-testing intervals of participants may have been increased during those weeks, resulting in somewhat lower viral loads at the time of inclusion in the study.

The largest differences in RT-PCR positivity percentages and Ag-RDTs’ performance were between confirmatory testers and individuals who attended the test-sites for other reasons. As expected, RT-PCR positivity percentages were close to 100% in the confirmatory testers and substantially lower (30% to 43%) in the ‘other reasons’ group. This agreed with test positivity percentages observed in national surveillance during the study. Logically, a higher proportion of confirmatory testers (93-95%) than other testers (76-77%) had viral loads above the used viral-load cut-off. Adding oropharyngeal to nasal sampling had a larger benefit in the other reasons group (10-18% increase in sensitivity) than in the confirmatory testers group (<1-4%) because the sensitivities were already very high in the latter group.

We found trends towards lower sensitivities in participants who had a previous SARS-CoV-2 infection (60-67%) compared to those who had not (72-83%), with non-overlapping confidence intervals for Flowflex with nasal self-sampling only. This trend was absent for MPBio with OP-N sampling. These findings should be interpreted with caution due to the larger uncertainty around these subgroup specific accuracy estimates. However, we observed similar trends for the BD-Veritor and SD-Biosensor Ag-RDTs with either professional or self-sampling in two previous studies.^5,7^ The lower sensitivities of Ag-RDTs in individuals with a prior infection may be explained by generally lower viral loads, with some individuals potentially carrying viral RNA in the absence of a productive infection (i.e. no viral antigen production). Another explanation might be that individuals with a prior infection have circulating anti-N-protein antibodies^20^ which might bind to the N protein produced during the new infection, and thereby hampering the binding of monoclonal antibodies against the N-protein in the test device. These effects might be test device-specific given the variability in the performance across the three AG-RDTs.^5^ We also found trends towards slightly higher sensitivities in participants who never received a COVID-19 vaccination for the MPBio and Clinitest Ag-RDTs with nasal self-sampling, but all confidence intervals overlapped with those who were vaccinated 1, 2 or 3 times and no differential impact was observed when combined OP-N sampling was applied. A detailed discussion of other subgroup analysis results can be found in supplementary material 4.

Our study is the only Ag-RDT diagnostic accuracy study thus far that was conducted during the emergence of Omicron, and that compared Ag-RDTs’ performances with nasal self-sampling versus combined OP-N self-sampling. Additional strengths include the large numbers of participants recruited at multiple test-sites, the low percentage of missing values, reference test sampling and Ag-RDT self-testing within a few hours, unsupervised self-testing conform the real-world context of self-testing, blinding of participants for the reference test result, blinding of laboratory staff for the Ag-RDT result, and the use of a viral load cut-off.

Our study also has some limitations. First, the sample size calculation was based on the primary analysis and diagnostic accuracy parameters are by definition less precise for stratified and weekly analyses. Second, we did not determine the virus lineage in individual samples but relied on the national pathogen surveillance data to estimate the weekly Omicron variant prevalence.^16^ This surveillance system includes approximately 2,000 random samples from positive samples across the country on a weekly basis. Since regional variations in the Netherlands are very small (data not shown), we are confident that Omicron’s share was over 90% in all test-sites from 12 January 2022 onwards. Third, the viral load cut-off that we used was the cut-off above which 95% of people with a positive RT-PCR test result had a positive virus culture in our similar previous study.^2^ Those experiments were done when the Alpha variant dominated and participants were mostly unvaccinated. However, we believe that this estimate is still more meaningful than using arbitrary Ct value cut-offs of 25 or 30, as is often done.^21,22^

We found that Ag-RDT performance with nasal self-sampling declined during the period that Omicron emerged. We also showed that Ag-RDT performance can be improved by adding oropharyngeal to nasal self-sampling. Therefore, after proper evaluation, Ag-RDT manufacturers may consider extending their instructions for use to include combined oropharyngeal and nasal self-sampling. Positive predictive values were high throughout our study, and symptomatic individuals can therefore rely on a positive Ag-RDT result irrespective of SARS-CoV-2 variant dominance or method of self-sampling. Negative predictive values were much lower. In mid-January 2022, the Dutch government advised all individuals with symptoms to do a self-test but advised vulnerable persons and those in close contacts with vulnerable persons to do RT-PCR tests at the PHS. In case of a negative self-test, individuals would be allowed to go to work or school. Our data show that this does reduce but not minimize transmission risks because of the likelihood of false-negative Ag-RDT results. As per national policy, we recommend that persons who test negative by self-test adhere to the general preventive measures such as applying hand hygiene, ensure indoor ventilation, and wearing mouth-nose masks in crowed places. In case of a positive self-test, self-isolation is required but confirmatory testing seems unnecessary in most situations if the infection pressure is high.

## Supporting information

STARD checklist

## Data Availability

Individual participant data collected during the study will be available, after deidentification of all participants. Data will be available to researchers who provide a methodologically sound proposal to achieve the aims in the approved proposal. Proposals should be directed to the corresponding author to gain access to the data. Data requestors will need to sign a data sharing agreement.

## Acknowledgements

We thank the participants, and study staff at the participating public health service test-sites, participating laboratories, the University Medical Center Utrecht, and RIVM for their contributions to the study. A special thanks to Esther Stiefelhagen, Renske Beekes, Sophie Neeleman, Eveline Westergaard, Roel Ensing, Wendy Mouthaan, and Timo Boelsums. Written permission was obtained from all five of them to list their names. ES, RB, SN, RE, WH, LB, and TB did not receive any compensation for their contributions.

## Contributors

KGMM initiated the study. ES, RPV, LH, IKV, WvdB, SDP, EL, MH, RM, CW, IV, CRSN-I, SvdH, JAJWK, JHHMvdW, and KGMM designed the study. ES, RPV, CRSN-I, and KGMM coordinated the study. WvdB, SDP, VFZ, LS, and MK were responsible for laboratory analyses and data processing. ES, RPV, and KGMM verified the underlying data. ES performed the statistical analysis and verified the underlying data in close collaboration with RPV and KGMM. ES, RPV, JHHMvdW, and KGMM drafted the first version of the manuscript. All authors critically read the manuscript and provided feedback. All authors approved the submission of the current version of the manuscript. The corresponding author attests that all listed authors meet authorship criteria and that no others meeting the criteria have been omitted.

## Ethical approval

Not required because the study was judged by the METC Utrecht to be outside the scope of the Dutch Medical Research Involving Human Subjects Act (protocol No 21-818 /C). All participants signed an informed consent form before any study procedure.

## Data sharing

The corresponding author (KGMM, the manuscript’s guarantor) affirms that the manuscript is an honest, accurate, and transparent account of the study being reported; that no important aspects of the study have been omitted; and that any discrepancies from the study as originally planned (and, if relevant, registered) have been explained.

## Additional information

The study protocol is available upon request by contacting Karel Moons at

## References

1. RIVM Centrum Infectieziektebestrijding. Status validatie SARS-CoV-2 antigeen sneltesten, 10 Mar 2021 [Dutch]. 2021. https://lci.rivm.nl/antigeensneltesten

2. Schuit E, Veldhuijzen IK, Venekamp RP, et al. Diagnostic accuracy of rapid antigen tests in asymptomatic and presymptomatic close contacts of individuals with confirmed SARS-CoV-2 infection: cross sectional study. BMJ. Jul 27 2021;374:n1676. doi:10.1136/bmj.n1676

3. Brummer LE, Katzenschlager S, Gaeddert M, et al. Accuracy of novel antigen rapid diagnostics for SARS-CoV-2: A living systematic review and meta-analysis. PLoS Med. Aug 2021;18(8):e1003735. doi:10.1371/journal.pmed.1003735

4. Scheiblauer H, Filomena A, Nitsche A, et al. Comparative sensitivity evaluation for 122 CE-marked rapid diagnostic tests for SARS-CoV-2 antigen, Germany, September 2020 to April 2021. Euro Surveill. Nov 2021;26(44)doi:10.2807/1560-7917.ES.2021.26.44.2100441

5. Venekamp RP, Veldhuijzen IK, Moons KGM, et al. Diagnostic accuracy of three prevailing rapid antigen tests for detection of SARS-CoV-2 infection in the general population: cross sectional study. medRxiv. 2021;

6. European Centre for Disease Prevention and Control (ECDC). Considerations on the use of self-tests for COVID-19 in the EU/EEA. 2021. ECDC Technical Report.

7. Schuit E, Venekamp RP, Veldhuijzen IK, et al. Accuracy and usability of saliva and nasal rapid antigen self-testing for detection of SARS-CoV-2 infection in the general population: a head-to-head comparison. medRxiv. 2021;doi:https://doi.org/10.1101/2021.12.08.21267452

8. Molenkamp R, Igloi Z. Evaluation of Antigen rapid test and PCR test to Omicron variant. 2021. https://www.erasmusmc.nl/-/media/erasmusmc/pdf/1-themaspecifiek/viroscience/2021-evaluation-omicron-in-pcr-and-ag-assays.pdf]

9. Deerain J, Druce J, Tran T, et al. Assessment of the Analytical Sensitivity of 10 Lateral Flow Devices against the SARS-CoV-2 Omicron Variant. J Clin Microbiol. Feb 16 2022;60(2):e0247921. doi:10.1128/jcm.02479-21

10. Osterman A, Badell I, Basara E, et al. Impaired detection of omicron by SARS-CoV-2 rapid antigen tests. Med Microbiol Immunol. Feb 20 2022;doi:10.1007/s00430-022-00730-z

11. de Michelena P, Torres I, Ramos-García A, et al. Real-life performance of a COVID-19 rapid antigen detection test targeting the SARS-CoV-2 nucleoprotein for diagnosis of COVID-19 due to the Omicron variant. medRxiv. 2022;doi:https://doi.org/10.1101/2022.02.02.22270295

12. Schrom J, Marquez C, Pilarowski G, et al. Direct Comparison of SARS-CoV-2 Nasal RT-PCR and Rapid Antigen Test (BinaxNOW™) at a Community Testing Site During an Omicron Surge. medRxiv. 2022;doi:https://doi.org/10.1101/2022.01.08.22268954

13. Bossuyt PM, Reitsma JB, Bruns DE, et al. STARD 2015: an updated list of essential items for reporting diagnostic accuracy studies. BMJ. Oct 28 2015;351:h5527. doi:10.1136/bmj.h5527

14. European Centre for Disease Prevention and Control (ECDC). Combined indicator: 14-day notification rate, testing rate and test positivity, updated 16 September 2021, weeks 35–36. Updated 16 September 2021. Accessed 7 December 2021, 2021. https://www.ecdc.europa.eu/en/publications-data/combined-indicator-week-36-2021

15. Rijksoverheid. Variants of the corona virus. Updated 2 December 2021. Accessed 2 December 2021, https://coronadashboard.rijksoverheid.nl/landelijk/varianten

16. RIVM Centrum Infectieziektebestrijding. Variants of the corona virus SARS-CoV-2 [Dutch]. Updated 30 November 2021. Accessed 2 December 2021, https://www.rivm.nl/coronavirus-covid-19/virus/varianten

17. RIVM Centrum Infectieziektebestrijding. Epidemiologische situatie van SARS-CoV-2 in Nederland [Dutch]. Updated 28 September 2021. Accessed 7 December 2021, 2021. https://www.rivm.nl/sites/default/files/2021-09/COVID-19_WebSite_rapport_wekelijks_20210928_1146_final.pdf

18. Klein JAF, Kruger LJ, Tobian F, et al. Head-to-head performance comparison of self-collected nasal versus professional-collected nasopharyngeal swab for a WHO-listed SARS-CoV-2 antigen-detecting rapid diagnostic test. Med Microbiol Immunol. Aug 2021;210(4):181–186. doi:10.1007/s00430-021-00710-9

19. Lindner AK, Nikolai O, Kausch F, et al. Head-to-head comparison of SARS-CoV-2 antigen-detecting rapid test with self-collected nasal swab versus professional-collected nasopharyngeal swab. Eur Respir J. Apr 2021;57(4)doi:10.1183/13993003.03961-2020

20. Allen N, Brady M, Carrion Martin AI, et al. Serological markers of SARS-CoV-2 infection; anti-nucleocapsid antibody positivity may not be the ideal marker of natural infection in vaccinated individuals. J Infect. Oct 2021;83(4):e9–e10. doi:10.1016/j.jinf.2021.08.012

21. Igloi Z, Velzing J, van Beek J, et al. Clinical Evaluation of Roche SD Biosensor Rapid Antigen Test for SARS-CoV-2 in Municipal Health Service Testing Site, the Netherlands. Emerg Infect Dis. May 2021;27(5):1323–1329. doi:10.3201/eid2705.204688

22. van Kampen JJA, van de Vijver D, Fraaij PLA, et al. Duration and key determinants of infectious virus shedding in hospitalized patients with coronavirus disease-2019 (COVID-19). Nat Commun. Jan 11 2021;12(1):267. doi:10.1038/s41467-020-20568-4

