## Supplementary Material for "Accuracy of COVID-19 self-tests with unsupervised nasal or nasal plus oropharyngeal self-sampling in symptomatic individuals in the Omicron period"

Table of contents

| | |
|---|---|
| **Supplementary material 1: short questionnaire after performing the self-test at home** | **3** |
| **Supplementary material 2: short questionnaire provided 10 days after test site visit** | **7** |
| **Supplementary material 3: Specimen collection, SARS-CoV-2 diagnostic testing, and SARS-CoV-2 virus culture procedures** | **9** |
| **Supplementary material 4: Discussion on subgroup effects based on gender and age.** | **11** |
| **Figure S1 Flow of study participants in the Delta-Omicron transition period** | **12** |
| **Figure S2 Sensitivities with 95% confidence intervals of the antigen rapid test-RT-PCR reference standard test comparisons after applying a viral load cut-off as a proxy for infectiousness, and stratified according to COVID-19 vaccination status, previous infection status, sex, and age, with nasal sampling for all three tests (top) and oropharyngeal plus nasal sampling for MPBio and Clinitest (bottom).** | **13** |
| **Figure S3 Sensitivities with 95% confidence intervals of the antigen rapid test-RT-PCR reference standard test comparisons stratified according to COVID-19 vaccination status, previous infection status, sex, and age, with nasal sampling, stratified by confirmatory testing (left) vs. other reason for testing (right).** | **14** |
| **Figure S4 Sensitivities with 95% confidence intervals of the antigen rapid test-RT-PCR reference standard test comparisons stratified according to COVID-19 vaccination status, previous infection status, sex, and age, with OP-N sampling, stratified by confirmatory testing (left) vs. other reason for testing (right).** | **15** |
| **Figure S5 Sensitivities with 95% confidence intervals of the antigen rapid test with nasal sampling-RT-PCR reference standard test comparison by week of inclusion stratified by confirmatory testing (black) or other reason for testing (red), before (left) and after application of a viral load cut-off (right).** | **16** |
| **Table S1 Baseline characteristics of the study population during the Delta-Omicron transition period, stratified by rapid antigen test** | **17** |
| **Table S2 2x2 tables for primary and secondary analyses of the RT-PCR reference test – Acon Labs Flowflex COVID-19 Antigen Home Test comparison – nasal sampling** | **19** |
| **Table S3 2x2 tables for primary and secondary analyses of the RT-PCR reference test – MP Biomedicals Rapid SARS-CoV-2 Antigen Test Card comparison – nasal sampling** | **20** |
| **Table S4 2x2 tables for primary and secondary analyses of the RT-PCR reference test – Siemens-Healthineers CLINITEST Rapid COVID-19 Antigen Test – nasal sampling** | **21** |
| **Table S5 2x2 tables for primary and secondary analyses of the RT-PCR reference test – MP Biomedicals Rapid SARS-CoV-2 Antigen Test Card comparison – nasal sampling** | **22** |

**Table S6 2x2 tables for primary and secondary analyses of the RT-PCR reference test – Siemens-Healthineers CLINITEST Rapid COVID-19 Antigen Test – nasal sampling** **23**

**Table S7 Diagnostic accuracy parameters for the three Ag-RDTs in symptomatic individuals in the Omicron era, stratified by reasons for testing.** **24**

**Table S8 User experience of the three Ag-RDTs** **25**

**Table S9 Follow-up information of participants that were tested again within 10 days after the initial test.** **27**

**Supplementary material 1: short questionnaire after performing both self-tests at home (translated from Dutch). Questions or items that were added for phase-2 (oropharyngeal + nasal sampling) are indicated in red.**

1. What was the result of the self-test you just performed for this study?
   ☐ Negative
   ☐ Positive
   ☐ Invalid
   ☐ I don't know: unable to perform the test
   ☐ I don't know: unable to read the test result
   ☐ Other, …………

2. Which sampling technique did you apply when performing the self-test, you just performed for this study?
   ☐ first oropharyngeal, then nasal sampling
   ☐ first nasal, then oropharyngeal sampling
   ☐ oropharyngeal sampling only
   ☐ nasal sampling only

3. What is your gender?
   ☐ Male
   ☐ Female
   ☐ Other (non-binary / genderqueer)

4. What is your age?
   ☐☐ years

5. What is the reason for testing?
   *Multiple answers possible*
   ☐ I have (had) Covid-19 like symptoms
   ☐ I am a close contact of a SARS-CoV-2 infected person
   - Date of last contact: ___ - ___ (day – month)

   I was notified because of:
   - The infected person is a household member □ yes / □ no
   - The infected person is a close contact (more than 15 minutes within 1.5 meters), but not a household member □ yes / □ no
   - Received notification by CoronaMelder app (English: Corona notification app) □ yes / □ no
   - Received notification by public health service (by phone or letter) □ yes / □ no
   - Received notification by SARS-CoV-2 infected person □ yes / □ no
   - Other, ………

   ☐ GP recommended a SARS-CoV-2 test
   ☐ Travelled to a high-pandemic country
   ☐ I performed a self-test and tested positive
   - What was the date of the positive self-test: ___ - ___ (day – month)

   ☐ None of the above, but…….

6. Did you receive a Covid-19 vaccination?
   ☐ No ***GO TO QUESTION 8***
   ☐ Yes

   If yes, how many vaccinations did you receive?
   ☐ One
   ☐ Two
   ☐ Three
   ☐ Four

   If yes, which vaccine did you receive (tip: this can be found in the CoronaCheck QR-code app under "International"/"Vaccination certificate")?
   Vaccine 1:
   ☐ Pfizer
   ☐ Moderna
   ☐ AstraZeneca

☐ Janssen
☐ Unknown/Other

Vaccine 2:
☐ Pfizer
☐ Moderna
☐ AstraZeneca
☐ Janssen
☐ Unknown/Other

Vaccine 3:
☐ Pfizer
☐ Moderna
☐ AstraZeneca
☐ Janssen
☐ Unknown/Other

Vaccine 4:
☐ Pfizer
☐ Moderna
☐ AstraZeneca
☐ Janssen
☐ Unknown/Other

7. What was the date of the last vaccination (tip: this can be found in the CoronaCheck QR-code app under "International"/"Vaccination certificate")?
   ___ - ___ - ___ (day – month – year)

8. Did you have a positive SARS-CoV-2 test previously?
   ☐ No ***GO TO QUESTION 9***
   ☐ Yes

   If yes, how often
   ☐☐ times

   If yes, how long ago?
   ☐ < 2 months
   ☐ 2-6 months
   ☐ 6-12 months
   ☐ >12 months

   If yes, how was the SARS-CoV-2 infection detected?
   ☐ via public health service
   ☐ via a commercial test sites ("testing for access" or "testing for travel")
   ☐ via a self-test
   ☐ Other (e.g., in another country, hospital, or at work): …………

   If yes, via a
   ☐ PCR test
   ☐ Rapid antigen test
   ☐ Unknown

   If yes, did this positive test occur after you had been vaccinated against COVID-19?
   ☐ No
   ☐ Yes, after:
      ☐ the first vaccination

☐ the second vaccination
☐ the third vaccination
☐ the fourth vaccination

9. At this moment, do you have any COVID-19 like symptoms?
☐ No ***GO TO QUESTION 12***
☐ Yes

10. What COVID-19 like symptoms do you currently have?
*Multiple answers possible*
☐ Common cold
☐ Shortness of breath
☐ Fever
☐ Coughing
☐ Loss of taste or smell
☐ Muscle ache
☐ I have other symptoms: ……….

11. What was the moment you first experienced these symptoms?
☐ Today
☐ Yesterday
☐ Two days ago
☐ Three or more days ago, at …/… (day/month)

12. Have you performed a self-test before?
☐ No ***GO TO QUESTION 13***
☐ Yes

If yes, how often?
☐☐ times

If yes, why?
☐ because of COVID-19 related symptoms
☐ because of an infected household member
☐ because of a close contact of an infected individual
☐ because I had to / it was recommended by my employer
☐ because I wanted to prevent me from potentially infected someone else because of the following situation: ………
☐ Other: ………

If yes, when did you perform your most recent self-test?
☐ less than 7 days ago
☐ 1-4 weeks ago
☐ 2-3 months ago
☐ 3-6 months ago
☐ more than 6 months ago

If yes, how did you obtain the self-test?
☐ received it from the government
☐ store-bought
☐ received it from school
☐ received it from work
☐ other: ………

13. Did you experience any difficulties when performing the self-test in this study?
☐ No ***GO TO QUESTION 14***
☐ Yes

If yes,

- ☐ I had a problem when performing the test:
  - ☐ Unsure how deep to place the swab up my throat
  - ☐ Fear of placing the swab too far up my throat
  - ☐ I had a gag reflex when placing the swab up my throat
  - ☐ Unsure how deep to place the swab up my nose
  - ☐ Fear of placing the swab too far up my nose
  - ☐ Spilled fluid
  - ☐ Other ………
- ☐ I had a problem interpreting the test:
  - ☐ I saw no control line
  - ☐ Liquid didn't travel through the cassette
  - ☐ Unclear whether the test was positive
  - ☐ Other ………

14. How difficult was it to perform oropharyngeal self-sampling, on a scale from 1 to 5?

    *Very easy 1 2 3 4 5 very hard*

15. How difficult was it to perform nasal self-sampling, on a scale from 1 to 5?

    *Very easy 1 2 3 4 5 very hard*

16. Do you agree with the following statements?

    The instruction for use of the self-test used in this study was clear
    *Agree In doubt Disagree*

    I am confident I performed the self-test used in this study correctly
    *Agree In doubt Disagree*

    I am confident I interpreted the self-test used in this study correctly
    *Agree In doubt Disagree*

    In the future, I will perform additional oropharyngeal sampling more often
    *Agree In doubt Disagree*

    I would rather perform nasal sampling only (instead of nasal plus oropharyngeal sampling)
    *Agree In doubt Disagree*

**Supplementary material 2: short questionnaire provided 10 days after test site visit (translated from Dutch). Questions or items that were added for phase-2 (oropharyngeal + nasal sampling) are indicated in red.**

1. Did you develop any COVID-19 like symptoms since your corona test at the public health service test site and the self-test performed for this study 10 days ago (see next question for list of possible symptoms)?
   - ☐ I had symptoms 10 days ago already and I experienced no new symptoms ***GO TO QUESTION 4***
   - ☐ I had symptoms 10 days ago already and I experienced new symptoms
   - ☐ I had no symptoms 10 days ago, but I experienced new symptoms
   - ☐ I had no symptoms 10 days ago and I experienced no new symptoms ***GO TO QUESTION 4***

2. What COVID-19 like symptoms did you have?
   *Multiple answers possible*
   - ☐ Common cold
   - ☐ Shortness of breath
   - ☐ Fever
   - ☐ Coughing
   - ☐ Loss of taste or smell
   - ☐ Muscle ache
   - ☐ I have other symptoms: ……….

3. Since when do you have these symptoms?
   … / … / … (day/month/year)

4. Did you perform a corona test or was a corona test performed in the last 10 days since your corona test at the public health service test site and the self-test performed for this study?
   - ☐ No ***THIS CONCLUDES THIS QUESTIONNAIRE***
   - ☐ Yes

   If yes, what type of test was performed?
   - ☐ Test at a public health service test site
     - ☐ PCR
     - ☐ (Rapid) antigen test
     - ☐ I don't know
   - ☐ Test at another test location (e.g., a commercial test site or at work)
     - ☐ PCR
     - ☐ (Rapid) antigen test
     - ☐ I don't know
   - ☐ Test at the hospital or general practitioner
     - ☐ PCR
     - ☐ (Rapid) antigen test
     - ☐ I don't know
   - ☐ Self-test
     - ☐ with nasal sampling
     - ☐ with nasal and oropharyngeal sampling
     - ☐ with oropharyngeal sampling
   - ☐ Other, ………

5. What is the reason for testing?
   *Multiple answers possible*
   - ☐ I have (had) Covid-19 like symptoms
   - ☐ I am a close contact of a corona infected person
   - Date of last contact: ___ - ___ (day – month)

   I was notified because of:
   - The infected person is a household member □ yes / □ no
   - The infected person is a close contact (more than 15 minutes within 1.5 meters), but not a household member □ yes / □ no
   - Received notification by CoronaMelder app (English: Corona notification app) □ yes / □ no
   - Received notification by public health service (by phone or letter) □ yes / □ no

- Received notification by a corona infected person □ yes / □ no
- Other, ………

☐ GP recommended a corona test
☐ Travelled to a high-pandemic country
☐ I performed a self-test and tested positive

- What was the date of the positive self-test: ___ - ___ (day – month)

☐ None of the above, but…….

6. Was at least one of these new corona tests positive?

☐ No
☐ Yes
☐ Unclear (result of self-test not clear clear), because……….

If yes, what was the date of this positive test: ___ - ___ (day – month)

**Supplementary material 3: Specimen collection, SARS-CoV-2 diagnostic testing, and SARS-CoV-2 virus culture procedures**

**Specimen collection and SARS-CoV-2 diagnostic testing procedures**

*West Brabant region, Microvida laboratory at Bravis hospital in Roosendaal; samples from Roosendaal*

One combined oropharyngeal and nasal swab (about 2.5 cm deep from the edge of the nostril) was taken per person at the Public Health Service test sites at the test sites in Roosendaal, placed in universal transport media (HiViralTM) with MagnaPure LC lysis- and binding buffer (Roche Diagnostics, The Netherlands), and transported to the Microvida laboratory in Roosendaal. RT-PCR was used as the reference standard test. RT-PCR was performed using the Cobas SARS-CoV-2 test on either the Cobas 6800 or the Cobas 8800 platform (Roche Diagnostics International, Rotkreuz, Switzerland). Cycle threshold (Ct) values for the SARS-CoV-2 E-gene were converted to viral load (genome copies/ml) using a previously established standard curve.[1,2] The assay was used in accordance with the manufacturer's instructions; amplification curves and Ct values were interpreted using the manufacturer's interpretation algorithms, which complied with the European in-vitro diagnostic devices directive.

*Central- and Northeast Brabant region, Sanquin Blood Supply Foundation in Amsterdam; samples from Tilburg (Ledeboer and Ringbaan-West)*

One superficial combined oropharyngeal and nasal swab (about 2.5 cm deep from the edge of the nostril) was taken per person at the Public Health Service test sites at the test sites in Tilburg, placed in universal transport media (HiViralTM) with MagnaPure LC lysis- and binding buffer (Roche Diagnostics, The Netherlands), and transported to the Sanquin Blood Supply Foundation in Amsterdam. RT-PCR was used as the reference standard test. RT-PCR was performed using the Cobas SARS-CoV-2 test on the Cobas 8800 platform (Roche Diagnostics International, Rotkreuz, Switzerland). Cycle threshold (Ct) values for the SARS-CoV-2 E-gene were converted to viral load (genome copies/ml) using a previously established standard curve.[1,2] The assay was used in accordance with the manufacturer's instructions; amplification curves and Ct values were interpreted using the manufacturer's interpretation algorithms, which complied with the European in-vitro diagnostic devices directive.

*Rotterdam region, Reinier Haga Medical Diagnostic center in Delft: samples from Rotterdam (Rotterdam-The Hague Airport)*

One deep combined OP-N swab (>5 cm deep from the edge of the nostril) swab was taken per person at the Public Health service test site in the Rotterdam region, placed directly in universal transport media (HiViralTM) with MagnaPure LC lysis- and binding buffer (Roche Diagnostics, The Netherlands), and transported to the Reinier Haga Medical Diagnostic Center. RT-PCR was used as the reference standard test. RT-PCR testing was performed in virus transport medium using the Cobas SARS-CoV-2 test on the Cobas 6800 platform. The assay was used in accordance with the manufacturer's instructions; amplification curves and cycle threshold values were interpreted using the manufacturer's interpretation algorithms, which complied with the European in-vitro diagnostic devices directive.

**Viral load calculation**

We used a viral load cut-off as a proxy of infectiousness of ≥5.2 log10 SARS-CoV-2 E-gene copies/mL, which was the viral load cut-off above which 95% of people with a positive RT-PCR test had a positive virus culture in a recent previous study by our group.[3] For that study, the Erasmus MC Viroscience laboratory created a standard curve by testing dilutions of a specific quantified E-gene transcript (primary standard) available from the European Virus Archive (EVAg[1]) with the RT-PCR described by Corman et al.[2] The relation between Ct value and number of E-gene copies/ml was determined by linear regression analysis. Based on this calibration curve a secondary standard derived from cell-cultured virus was prepared and quantified. Serial dilutions of this secondary standard were tested by RT-PCR to prepare a secondary standard curve by linear regression.

In that previous study, we determined whether the Cobas PCR platform at the Microvida laboratory provided comparable data to the Cobas PCR platform at the Erasmus Medical Center Viroscience laboratory by having both laboratories test the same SARS-CoV-2 viral load panel obtained from the National Public Health Institute (RIVM). The Ct values generated in the two laboratories corresponded well. Therefore, we concluded that the same formula to convert Ct values into viral loads (copies/ml) could be used for the West-Brabant and Rotterdam laboratories. This formula was $62.5 * e^{\frac{43.1-Ct}{1.607}}$. Similarly, and as an extension, the most recent (October 2021) SARS-CoV-2 viral load panel (LEQA4) obtained from the National Public Health Institute (RIVM) was tested at the Sanquin Blood Supply Foundation and the Reinier Haga Medical Diagnostic Center. The Cobas platforms at all laboratories generated Ct values that were comparable. Therefore, we concluded that the same formula could be applied to all three laboratories used in the current study.

The three laboratories used different medium/reagent volumes per swab. For each, a factor $X$ was calculated based on the swab volume, dilution, and total sample volume, to allow alignment across samples with respect to the viral load in the Erasmus MC Viroscience laboratory standard. For example, if the sample volume is equal (3 ml in Tilburg compared to 3 ml in Rotterdam), but the dilution factor is higher, e.g., by the addition of lysis buffer (3 in Tilburg compared to 2 in Rotterdam), the effective viral sample would be lower than in the Rotterdam standard, thereby underestimating the actual viral load when using the above formula. To account for this, a factor $X$ < 1 was used, here 6/9. If dilution was lower, a factor $X$ > 1 was used. The conversion formula used therefore was $62.5 * e^{\frac{43.1-Ct}{1.607}} / X$.

**Self-tests**

*Acon Labs Flowflex COVID-19 Antigen Home Test*

The Acon Labs Flowflex COVID-19 Antigen Home Test is a CE-marked Ag-RDT. According to the instructions for use, a sample should be taken by placing a nasal swab less than 2.5 cm from the nostril edge in the nose and twisting it 5 times, once in each nostril. Afterwards the nose swab had to be placed in a test tube that was filled with buffer fluid and twisted for 30 seconds. Then the swab had to be twisted 5 times during which the test tube had to be squeezed lightly. While squeezing the tube, the swab could be removed from the tube, while leaving all the test fluid in the tube. After placing a cap on the tube, the tube had to be moved from left to right. Then, 4 droplets of the solvent had to be added to the testing cassette. After 15 minutes, but no later than 30 minutes, the participant had to read the test result visually from the testing cassette.

*MP Biomedicals Rapid SARS-CoV-2 Antigen Test Card*

The MP Biomedicals Rapid SARS-CoV-2 Antigen Test Card is a CE-marked Ag-RDT. According to the instructions for use, first, extraction fluid had to be placed into a test tube. A sample should be taken by placing a nasal swab at least 2.5 cm deep in the nose and twisting it 3-4 times, once in each nostril. Afterwards the nose swab had to be placed in a test tube that was filled by buffer fluid and twisted 3-5 times, after which the swab had to be left in the test tube for 1 minute. While squeezing the tube, the swab could be removed from the tube, while leaving all the test fluid in the tube. Then, after placing a cap on the tube, 3 droplets of the solvent had to be added to the testing cassette. After 15 to 20 minutes, but no later than 20 minutes, the participant had to read the test result visually from the testing cassette.

*Siemens-Healthineers CLINITEST Rapid COVID-19 Antigen Test*

The Siemens-Healthineers CLINITEST Rapid COVID-19 Antigen Test is a CE-marked Ag-RDT. According to the instructions for use, a sample should be taken by placing a nasal swab 2-4 cm from the nostril edge in the nose and twisting it 5 times, once in each nostril. Afterwards the nose swab had to be placed in a test tube that was filled with buffer fluid and twisted for 5 times, while touching the bottom and sidewalls of the test tube, after which the swab had to be left in the test tube for 1 minute. After squeezing the tube several times, the swab could be removed from the tube, while leaving all the test fluid in the tube. Then, after placing a cap on the tube, 4 droplets of the solvent had to be added to the testing cassette. After 15 minutes, the participant had to read the test result visually from the testing cassette.

**References**


1. Archive EV. Wuhan coronavirus 2019 E gene control. Accessed June 8 2021, 2021. https://www.european-virus-archive.com/nucleic-acid/wuhan-coronavirus-2019-e-gene-control
2. Corman VM, Landt O, Kaiser M, et al. Detection of 2019 novel coronavirus (2019-nCoV) by real-time RT-PCR. *Euro Surveill*. Jan 2020;25(3)doi:10.2807/1560-7917.ES.2020.25.3.2000045
3. Schuit E, Veldhuijzen IK, Venekamp RP, et al. Diagnostic accuracy of rapid antigen tests in asymptomatic and presymptomatic close contacts of individuals with confirmed SARS-CoV-2 infection: cross sectional study. *BMJ*. Jul 27 2021;374:n1676. doi:10.1136/bmj.n1676

**Supplementary material 4: Discussion on subgroup effects based on gender and age.**

We found differences in sensitivity between men and women (Flowflex and Clinitest with nasal sampling) and those aged 16-40 and >40 (MPBio with nasal sampling). To further assess these differences cross-tabulation was performed of gender and age groups with other subgroup characteristics for which we found distinct differences in sensitivity: confirmatory testing as reason for testing (yes [higher sensitivity] vs. no), vaccination status (no [higher sensitivity] vs. one or more vaccinations) and having had a previous SARS-CoV-2 infection (yes vs. no [higher sensitivity]). Cross-tabulation showed that women in the Flowflex study group less often visited the test site for confirmatory testing (42% vs. 49%) and more often had had a previous SARS-CoV-2 infection (28% vs. 18%) than men, both of which may explain the lower overall sensitivity in women vs. men with Flowflex. In the Clinitest study group, differences were smaller between women and men for these subgroup characteristics but were consistently favouring men, which may contribute to the lower sensitivity seen in women. In the MPBio study group, participants aged 40 years and older were less often confirmatory testers (25.1% vs. 32.5%), and less often not vaccinated (3.9% vs. 11.7%) than participants aged 16 to 40 years old, both of which may have contributed to the lower sensitivity in the older participants. Apparently, underrepresentation of confirmatory testers and non-vaccinators in older participants was not counterbalanced by the fact that older participants less often had had a SARS-CoV-2 infection (19.3% vs. 25.1%) than younger participants. To conclude, we do not expect the diagnostic accuracy of the three Ag-RDTs truly differs according to gender or age.

**Figure S1 Flow of study participants in the Delta-Omicron transition period**

See separate file: Figure S1.pdf

**Figure S2 Sensitivities with 95% confidence intervals of the antigen rapid test-RT-PCR reference standard test comparisons after applying a viral load cut-off as a proxy for infectiousness, and stratified according to COVID-19 vaccination status, previous infection status, sex, and age, with nasal sampling for all three tests (top) and oropharyngeal plus nasal sampling for MPBio and Clinitest (bottom). The vertical line indicates the sensitivity of the Ag-RDT in the respective overall study population before application of a viral load cut-off, and the number of positive RT-PCR tests out of the total or subgroup between parentheses.**

See separate file: Figure S2.pdf

**Figure S3 Sensitivities with 95% confidence intervals of the antigen rapid test-RT-PCR reference standard test comparisons stratified according to COVID-19 vaccination status, previous infection status, sex, and age, with nasal sampling, stratified by confirmatory testing (left) vs. other reason for testing (right). The vertical line indicates the sensitivity of the Ag-RDT in the respective overall study population, and the number of positive RT-PCR tests out of the total or subgroup between parentheses.**

See separate file: Figure S3.pdf

**Figure S4 Sensitivities with 95% confidence intervals of the antigen rapid test-RT-PCR reference standard test comparisons stratified according to COVID-19 vaccination status, previous infection status, sex, and age, with OP-N sampling, stratified by confirmatory testing (left) vs. other reason for testing (right). The vertical line indicates the sensitivity of the Ag-RDT in the respective overall study population, and the number of positive RT-PCR tests out of the total or subgroup between parentheses.**

See separate file: Figure S4.pdf

**Figure S5 Sensitivities with 95% confidence intervals of the antigen rapid test with nasal sampling-RT-PCR reference standard test comparison by week of inclusion stratified by confirmatory testing (black) or other reason for testing (red), before (left) and after application of a viral load cut-off (right). The histogram indicates the percentage of the Omicron variant according to the national pathogen surveillance, while the numbers indicate the number of participants included in each week, stratified by reason for testing.**

See separate file: Figure S5.pdf

**Table S1 Baseline characteristics of the study population during the Delta-Omicron transition period, stratified by rapid antigen test**

| Test result available from the reference test and… | Flowflex | MPBio | Clinitest |
|---|---|---|---|
| Location | Rotterdam | Tilburg | Roosendaal |
| Inclusion period | Dec 22 – Jan 11 | Dec 24 – Jan 11 | Dec 21 – Jan 11 |
| Sample size | N = 1303 | N = 866 | N = 853 |
| Age [years], mean (SD) | 37 (13) | 38 (15) | 41 (15) |
| Range (min-max) | 16-80 | 16-79 | 16-79 |
| Sex, female, No. (%) | 718 (55.1) | 567 (64.0) | 524 (61.4) |
| Reason for testing, No. (%)[4] | | | |
| Positive self-test | 304 (23.3) | 230 (26.0) | 228 (26.7) |
| Symptoms | 930 (71.4) | 592 (66.8) | 590 (69.2) |
| Close contact[1] | 183 (14.0) | 164 (18.5) | 130 (15.2) |
| Other reason | 106 (8.1) | 73 (8.2) | 68 (8.0) |
| Vaccination status | | | |
| Not vaccinated | 109 (8.4) | 61 (6.9) | 81 (9.5) |
| Vaccinated with at least one dose, No. (%) | 1194 (91.6) | 825 (93.1) | 772 (90.5) |
| Number of vaccinations received, No. (%)[2] | | | |
| 1 | 158 (13.2) | 106 (12.8) | 109 (14.1) |
| 2 | 858 (71.9) | 533 (64.6) | 546 (70.7) |
| 3 | 177 (14.8) | 186 (22.5) | 117 (15.2) |
| 4 | 1 (0.1) | 0 (0) | 0 (0) |
| Unknown | 0 (0) | 0 (0) | 0 (0) |
| Type of initial vaccine, No. (%)[2] | | | |
| Pfizer | 924 (77.4) | 539 (65.3) | 483 (62.6) |
| Moderna | 97 (8.1) | 101 (12.2) | 121 (15.7) |
| Astra Zeneca | 76 (6.4) | 104 (12.6) | 101 (13.1) |
| Janssen | 91 (7.6) | 79 (9.6) | 63 (8.2) |
| Unknown/Other | 6 (0.5) | 2 (0.2) | 4 (0.5) |
| Type of booster vaccine, No. (%)[2] | | | |
| Pfizer | 131 (11.0) | 116 (14.1) | 82 (10.6) |
| Moderna | 64 (5.4) | 75 (9.1) | 42 (5.4) |
| No booster received | 993 (83.2) | 621 (75.3) | 641 (83.0) |
| Unknown | 6 (0.5) | 13 (1.6) | 7 (0.9) |
| At least one prior SARS-CoV-2 infection, No. (%) | 317 (24.4) | 161 (18.2) | 156 (18.3) |
| Less than 2 months ago | 34 (10.7) | 16 (9.9) | 15 (9.6) |

| | | | |
|---|---|---|---|
| 2 to 6 months ago | 56 (17.7) | 20 (12.4) | 15 (9.6) |
| 6 to 12 months ago | 113 (35.6) | 69 (42.9) | 52 (33.3) |
| More than 12 months ago | 114 (36.0) | 56 (34.8) | 73 (46.8) |
| Unknown | 0 (0) | 0 (0) | 1 (0.6) |
| Symptom onset, No. (%)[3] | | | |
| On day of sampling | 148 (11.4) | 69 (7.8) | 58 (6.8) |
| A day before sampling | 492 (37.8) | 285 (32.2) | 313 (36.7) |
| Two days before sampling | 341 (26.2) | 289 (32.6) | 249 (29.2) |
| Three or more days before sampling | 321 (24.6) | 243 (27.4) | 232 (27.2) |
| Unknown | 1 (0.1) | 0 (0) | 1 (0.1) |
| Type of symptoms (self-reported), No. (%)[3,4] | | | |
| Common cold | 1113 (85.4) | 771 (87.0) | 730 (85.6) |
| Shortness of breath | 172 (13.2) | 148 (16.7) | 145 (17.0) |
| Fever | 257 (19.7) | 194 (21.9) | 177 (20.8) |
| Coughing | 592 (45.4) | 430 (48.5) | 440 (51.6) |
| Loss of taste or smell | 60 (4.6) | 40 (4.5) | 44 (5.2) |
| Muscle ache | 210 (16.1) | 181 (20.4) | 168 (19.7) |
| Other symptoms | 130 (10.0) | 117 (13.2) | 98 (11.5) |
| Previous experience with using self-tests, No. (%) | 1236 (94.9) | 851 (96.0) | 795 (93.5) |
| Performed last self-test, No. (%)[2] | | | |
| Less than 7 days ago | 1011 (81.8) | 723 (85.0) | 644 (81.0) |
| 1 to 4 weeks ago | 144 (11.7) | 77 (9.0) | 96 (12.1) |
| More than 1 months ago | 77 (6.2) | 50 (5.9) | 55 (6.9) |
| Unknown | 4 (0.3) | 1 (0.1) | 0 (0) |
| Number of performed self-tests, No. (%) | | | |
| 1-3 | 299 (24.2) | 211 (24.8) | 251 (31.6) |
| 4-6 | 362 (29.3) | 254 (29.9) | 248 (31.2) |
| 7-10 | 294 (23.8) | 189 (22.2) | 168 (21.1) |
| >10 | 279 (22.6) | 196 (23.1) | 128 (12.6) |

SD=standard deviation.

[1] In the Netherlands, individuals are notified of a close contact by the Dutch public health service test-and-trace program, and/or the Dutch contact tracing mobile phone application (the CoronaMelder app) and/or an individual with a confirmed SARS-CoV-2 infection (index case).

[2] percentage calculated as proportion of those vaccinated, or those that had previous experience in performing a self-test

[3] Percentage calculated as proportion of those with symptoms at time of sampling

[4] Totals add up to a number higher than the number of participants, and individuals with symptoms at the time of sampling because individuals could report more than one reason for testing, or symptom, respectively.

**Table S2** 2x2 tables for primary and secondary analyses of the RT-PCR reference test – Acon Labs Flowflex COVID-19 Antigen Home Test comparison – nasal sampling

| Analysis | Subgroup | | | | |
|---|---|---|---|---|---|
| **Primary** | | | Ref + | Ref - | Total |
| | | Test + | 323 | 6 | 329 |
| | | Test - | 86 | 205 | 291 |
| | | Total | 409 | 211 | 620 |
| **Secondary (stratified):** | | | | | |
| Infectiousness viral load cut-off¶ | | | Ref + | Ref - | Total |
| | | Test + | 309 | 20 | 329 |
| | | Test - | 52 | 239 | 291 |
| | | Total | 361 | 259 | 620 |
| Vaccinated (at least at least once): | Yes | | Ref + | Ref - | Total |
| | | Test + | 275 | 6 | 281 |
| | | Test - | 75 | 189 | 264 |
| | | Total | 350 | 195 | 545 |
| | No | | Ref + | Ref - | Total |
| | | Test + | 48 | 0 | 48 |
| | | Test - | 11 | 16 | 27 |
| | | Total | 59 | 16 | 75 |
| Previous SARS-CoV-2 infection: | Yes | | Ref + | Ref - | Total |
| | | Test + | 63 | 1 | 64 |
| | | Test - | 31 | 53 | 84 |
| | | Total | 94 | 54 | 148 |
| | No | | Ref + | Ref - | Total |
| | | Test + | 260 | 5 | 265 |
| | | Test - | 55 | 152 | 207 |
| | | Total | 315 | 157 | 472 |
| Sex: | Female | | Ref + | Ref - | Total |
| | | Test + | 171 | 2 | 173 |
| | | Test - | 57 | 127 | 184 |
| | | Total | 228 | 129 | 357 |
| | Male | | Ref + | Ref - | Total |
| | | Test + | 149 | 4 | 153 |
| | | Test - | 29 | 78 | 107 |
| | | Total | 178 | 82 | 260 |
| Age [years] | ≥16 to ≤40 | | Ref + | Ref - | Total |
| | | Test + | 201 | 5 | 206 |
| | | Test - | 53 | 126 | 179 |
| | | Total | 254 | 131 | 385 |
| | >40 | | Ref + | Ref - | Total |
| | | Test + | 122 | 1 | 123 |
| | | Test - | 33 | 79 | 112 |
| | | Total | 155 | 80 | 235 |
| Reasons for testing was positive self-test: | Yes | | Ref + | Ref - | Total |
| | | Test + | 247 | 3 | 250 |
| | | Test - | 17 | 12 | 29 |
| | | Total | 264 | 15 | 279 |
| | No | | Ref + | Ref - | Total |
| | | Test + | 76 | 3 | 79 |
| | | Test - | 69 | 193 | 262 |
| | | Total | 145 | 196 | 341 |

Ref = RT-PCR reference standard test; Test = Flowflex

¶Viral load cut-off for infectiousness, defined as viral load above which 95% of people with a positive RT-PCR test result had a positive viral culture[1] was 5.2 log10 SARS-CoV-2 E-gene copies/mL

**Table S3** 2x2 tables for primary and secondary analyses of the RT-PCR reference test – MP Biomedicals Rapid SARS-CoV-2 Antigen Test Card comparison – nasal sampling

| Analysis | Subgroup | | | | |
|---|---|---|---|---|---|
| **Primary** | | | Ref + | Ref - | Total |
| | | Test + | 276 | 5 | 281 |
| | | Test - | 119 | 420 | 539 |
| | | Total | 395 | 425 | 820 |
| **Secondary (stratified):** | | | | | |
| Infectiousness viral load cut-off¶ | | | Ref + | Ref - | Total |
| | | Test + | 267 | 14 | 281 |
| | | Test - | 73 | 465 | 538 |
| | | Total | 340 | 479 | 819 |
| Vaccinated (at least at least once): | Yes | | Ref + | Ref - | Total |
| | | Test + | 245 | 3 | 248 |
| | | Test - | 114 | 390 | 504 |
| | | Total | 359 | 393 | 752 |
| | No | | Ref + | Ref - | Total |
| | | Test + | 31 | 2 | 33 |
| | | Test - | 5 | 30 | 35 |
| | | Total | 36 | 32 | 68 |
| Previous SARS-CoV-2 infection: | Yes | | Ref + | Ref - | Total |
| | | Test + | 40 | 0 | 40 |
| | | Test - | 26 | 119 | 145 |
| | | Total | 66 | 119 | 185 |
| | No | | Ref + | Ref - | Total |
| | | Test + | 236 | 5 | 241 |
| | | Test - | 93 | 300 | 393 |
| | | Total | 329 | 305 | 634 |
| Sex: | Female | | Ref + | Ref - | Total |
| | | Test + | 166 | 3 | 169 |
| | | Test - | 70 | 285 | 355 |
| | | Total | 236 | 288 | 524 |
| | Male | | Ref + | Ref - | Total |
| | | Test + | 110 | 2 | 112 |
| | | Test - | 49 | 134 | 183 |
| | | Total | 159 | 136 | 295 |
| Age [years] | ≥16 to ≤40 | | Ref + | Ref - | Total |
| | | Test + | 177 | 2 | 179 |
| | | Test - | 58 | 238 | 296 |
| | | Total | 235 | 240 | 475 |
| | >40 | | Ref + | Ref - | Total |
| | | Test + | 99 | 3 | 102 |
| | | Test - | 61 | 182 | 243 |
| | | Total | 160 | 185 | 345 |
| Reasons for testing was positive self-test: | Yes | | Ref + | Ref - | Total |
| | | Test + | 189 | 2 | 191 |
| | | Test - | 37 | 11 | 48 |
| | | Total | 226 | 13 | 239 |
| | No | | Ref + | Ref - | Total |
| | | Test + | 87 | 3 | 90 |
| | | Test - | 82 | 409 | 491 |
| | | Total | 169 | 412 | 581 |

Ref = RT-PCR reference standard test; Test = MPBio

¶Viral load cut-off for infectiousness, defined as viral load above which 95% of people with a positive RT-PCR test result had a positive viral culture[1] was 5.2 log10 SARS-CoV-2 E-gene copies/mL.

**Table S4** 2x2 tables for primary and secondary analyses of the RT-PCR reference test – Siemens-Healthineers CLINITEST Rapid COVID-19 Antigen Test – nasal sampling

| Analysis | Subgroup | | | | |
|---|---|---|---|---|---|
| **Primary** | | | Ref + | Ref - | Total |
| | | Test + | 301 | 2 | 303 |
| | | Test - | 128 | 295 | 423 |
| | | Total | 429 | 297 | 726 |
| **Secondary (stratified):** | | | | | |
| Infectiousness viral load cut-off¶ | | | Ref + | Ref - | Total |
| | | Test + | 285 | 9 | 294 |
| | | Test - | 85 | 332 | 417 |
| | | Total | 370 | 341 | 711 |
| Vaccinated (at least at least once): | Yes | | Ref + | Ref - | Total |
| | | Test + | 255 | 0 | 255 |
| | | Test - | 117 | 263 | 380 |
| | | Total | 372 | 263 | 635 |
| | No | | Ref + | Ref - | Total |
| | | Test + | 46 | 2 | 48 |
| | | Test - | 11 | 31 | 42 |
| | | Total | 57 | 33 | 90 |
| Previous SARS-CoV-2 infection: | Yes | | Ref + | Ref - | Total |
| | | Test + | 38 | 2 | 40 |
| | | Test - | 25 | 62 | 87 |
| | | Total | 63 | 64 | 127 |
| | No | | Ref + | Ref - | Total |
| | | Test + | 263 | 0 | 263 |
| | | Test - | 103 | 233 | 336 |
| | | Total | 366 | 233 | 599 |
| Sex: | Female | | Ref + | Ref - | Total |
| | | Test + | 169 | 2 | 171 |
| | | Test - | 90 | 188 | 278 |
| | | Total | 259 | 190 | 449 |
| | Male | | Ref + | Ref - | Total |
| | | Test + | 131 | 0 | 131 |
| | | Test - | 38 | 107 | 145 |
| | | Total | 169 | 107 | 276 |
| Age [years] | ≥16 to ≤40 | | Ref + | Ref - | Total |
| | | Test + | 170 | 1 | 171 |
| | | Test - | 69 | 145 | 214 |
| | | Total | 239 | 146 | 385 |
| | >40 | | Ref + | Ref - | Total |
| | | Test + | 131 | 1 | 132 |
| | | Test - | 59 | 150 | 209 |
| | | Total | 190 | 151 | 341 |
| Reasons for testing was positive self-test: | Yes | | Ref + | Ref - | Total |
| | | Test + | 210 | 1 | 211 |
| | | Test - | 35 | 11 | 46 |
| | | Total | 245 | 12 | 257 |
| | No | | Ref + | Ref - | Total |
| | | Test + | 91 | 1 | 92 |
| | | Test - | 93 | 284 | 377 |
| | | Total | 184 | 285 | 469 |

Ref = RT-PCR reference standard test; Test = Clinitest

¶Viral load cut-off for infectiousness, defined as viral load above which 95% of people with a positive RT-PCR test result had a positive viral culture[1] was 5.2 log10 SARS-CoV-2 E-gene copies/mL.

**Table S5** 2x2 tables for primary and secondary analyses of the RT-PCR reference test – MP Biomedicals Rapid SARS-CoV-2 Antigen Test Card comparison – oropharyngeal-nasal sampling

| Analysis | Subgroup | | | | |
|---|---|---|---|---|---|
| **Primary** | | | Ref + | Ref - | Total |
| | | Test + | 303 | 4 | 307 |
| | | Test - | 62 | 175 | 237 |
| | | Total | 365 | 179 | 544 |
| **Secondary (stratified):** | | | | | |
| Infectiousness viral load cut-off¶ | | | Ref + | Ref - | Total |
| | | Test + | 283 | 24 | 307 |
| | | Test - | 32 | 205 | 237 |
| | | Total | 315 | 229 | 544 |
| Vaccinated (at least at least once): | Yes | | Ref + | Ref - | Total |
| | | Test + | 269 | 4 | 273 |
| | | Test - | 57 | 158 | 215 |
| | | Total | 326 | 162 | 488 |
| | No | | Ref + | Ref - | Total |
| | | Test + | 34 | 0 | 34 |
| | | Test - | 5 | 17 | 22 |
| | | Total | 39 | 17 | 56 |
| Previous SARS-CoV-2 infection: | Yes | | Ref + | Ref - | Total |
| | | Test + | 75 | 1 | 76 |
| | | Test - | 13 | 46 | 59 |
| | | Total | 88 | 47 | 135 |
| | No | | Ref + | Ref - | Total |
| | | Test + | 228 | 3 | 231 |
| | | Test - | 49 | 129 | 178 |
| | | Total | 277 | 132 | 409 |
| Sex: | Female | | Ref + | Ref - | Total |
| | | Test + | 202 | 3 | 205 |
| | | Test - | 43 | 128 | 171 |
| | | Total | 245 | 131 | 376 |
| | Male | | Ref + | Ref - | Total |
| | | Test + | 99 | 1 | 100 |
| | | Test - | 19 | 47 | 66 |
| | | Total | 118 | 48 | 166 |
| Age [years] | ≥16 to ≤40 | | Ref + | Ref - | Total |
| | | Test + | 189 | 3 | 192 |
| | | Test - | 40 | 104 | 144 |
| | | Total | 229 | 107 | 336 |
| | >40 | | Ref + | Ref - | Total |
| | | Test + | 114 | 1 | 115 |
| | | Test - | 22 | 71 | 93 |
| | | Total | 136 | 72 | 208 |
| Reasons for testing was positive self-test: | Yes | | Ref + | Ref - | Total |
| | | Test + | 242 | 1 | 243 |
| | | Test - | 35 | 10 | 45 |
| | | Total | 277 | 11 | 288 |
| | No | | Ref + | Ref - | Total |
| | | Test + | 61 | 3 | 64 |
| | | Test - | 27 | 165 | 192 |
| | | Total | 88 | 168 | 256 |

Ref = RT-PCR reference standard test; Test = MPBio

¶Viral load cut-off for infectiousness, defined as viral load above which 95% of people with a positive RT-PCR test result had a positive viral culture[1] was 5.2 log10 SARS-CoV-2 E-gene copies/mL.

**Table S6** 2x2 tables for primary and secondary analyses of the RT-PCR reference test – Siemens-Healthineers CLINITEST Rapid COVID-19 Antigen Test – oropharyngeal-nasal sampling

| **Analysis** | **Subgroup** | | | | |
|---|---|---|---|---|---|
| **Primary** | | | Ref + | Ref - | Total |
| | | Test + | 326 | 7 | 333 |
| | | Test - | 96 | 224 | 320 |
| | | Total | 422 | 231 | 653 |
| **Secondary (stratified):** | | | | | |
| Infectiousness viral load cut-off¶ | | | Ref + | Ref - | Total |
| | | Test + | 313 | 20 | 333 |
| | | Test - | 61 | 259 | 320 |
| | | Total | 374 | 279 | 653 |
| Vaccinated (at least at least once): | Yes | | Ref + | Ref - | Total |
| | | Test + | 292 | 4 | 296 |
| | | Test - | 87 | 203 | 290 |
| | | Total | 379 | 207 | 586 |
| | No | | Ref + | Ref - | Total |
| | | Test + | 34 | 2 | 36 |
| | | Test - | 9 | 21 | 30 |
| | | Total | 43 | 23 | 66 |
| Previous SARS-CoV-2 infection: | Yes | | Ref + | Ref - | Total |
| | | Test + | 40 | 3 | 43 |
| | | Test - | 20 | 58 | 78 |
| | | Total | 60 | 61 | 121 |
| | No | | Ref + | Ref - | Total |
| | | Test + | 286 | 4 | 290 |
| | | Test - | 76 | 166 | 242 |
| | | Total | 362 | 170 | 532 |
| Sex: | Female | | Ref + | Ref - | Total |
| | | Test + | 197 | 6 | 203 |
| | | Test - | 66 | 168 | 234 |
| | | Total | 263 | 174 | 437 |
| | Male | | Ref + | Ref - | Total |
| | | Test + | 127 | 1 | 128 |
| | | Test - | 30 | 55 | 85 |
| | | Total | 157 | 56 | 213 |
| Age [years] | ≥16 to ≤40 | | Ref + | Ref - | Total |
| | | Test + | 174 | 4 | 178 |
| | | Test - | 46 | 115 | 161 |
| | | Total | 220 | 119 | 339 |
| | >40 | | Ref + | Ref - | Total |
| | | Test + | 152 | 3 | 155 |
| | | Test - | 50 | 108 | 158 |
| | | Total | 202 | 111 | 313 |
| Reasons for testing was positive self-test: | Yes | | Ref + | Ref - | Total |
| | | Test + | 241 | 2 | 243 |
| | | Test - | 39 | 8 | 47 |
| | | Total | 208 | 10 | 290 |
| | No | | Ref + | Ref - | Total |
| | | Test + | 85 | 5 | 90 |
| | | Test - | 57 | 216 | 273 |
| | | Total | 142 | 221 | 363 |

Ref = RT-PCR reference standard test; Test = Clinitest

¶Viral load cut-off for infectiousness, defined as viral load above which 95% of people with a positive RT-PCR test result had a positive viral culture[1] was 5.2 log10 SARS-CoV-2 E-gene copies/mL.

**Table S7 Diagnostic accuracy parameters for the three Ag-RDTs in symptomatic individuals in the Omicron era, stratified by reasons for testing. Values are percentages (95% confidence interval) unless stated otherwise. Categories for reasons of testing are not mutually exclusive; participants could report multiple reasons for testing. If a participant indicated multiple reasons, he or she was included in the analysis of both categories.**

| Analysis | Sampling | No. | Prevalence* [%] | Sensitivity [%] (95%CI) | Specificity [%] (95%CI) | PPV [%] (95%CI) | NPV [%] (95%CI) |
|---|---|---|---|---|---|---|---|
| **Flowflex** | | | | | | | |
| Reason for testing: | | | | | | | |
| Positive self-test | N | 279 | 94.6 | 93.6 (89.9-96.2) | 80.0 (51.9-95.7) | 98.8 (96.5-99.8) | 41.4 (23.5-61.1) |
| Symptoms | N | 405 | 59.8 | 77.7 (71.9-82.8) | 96.9 (93.0-99.0) | 97.4 (94.1-99.2) | 74.5 (68.1-80.2) |
| Close contact | N | 72 | 51.4 | 51.4 (34.4-68.1) | 100 (90.0-100) | 100 (82.4-100) | 66.0 (51.7-78.5) |
| Other reason | N | 49 | 69.4 | 61.8 (43.6-77.8) | 100 (78.2-100) | 100 (83.9-100) | 53.6 (33.9-72.5) |
| **MPBio** | | | | | | | |
| Reason for testing: | | | | | | | |
| Positive self-test | N | 239 | 94.6 | 83.6 (78.1-88.2) | 84.6 (54.6-98.1) | 99.0 (96.3-99.9) | 22.9 (12.0-37.3) |
| | OP-N | 288 | 96.2 | 87.4 (82.9-91.0) | 90.9 (58.7-99.8) | 99.6 (97.7-100) | 22.2 (11.2-37.1) |
| Symptoms | N | 510 | 41.2 | 66.7 (59.9-73.0) | 98.7 (96.6-99.6) | 97.2 (93.0-99.2) | 80.9 (76.5-84.8) |
| | OP-N | 355 | 60.8 | 79.6 (73.6-84.8) | 87.1 (92.8-99.2) | 97.7 (94.3-99.4) | 75.4 (68.4-81.5) |
| Close contact | N | 170 | 37.1 | 60.3 (47.2-72.4) | 99.1 (94.9-100) | 97.4 (86.5-99.9) | 80.9 (73.1-87.3) |
| | OP-N | 58 | 37.9 | 77.3 (54.6-92.2) | 100 (90.3-100) | 100 (80.5-100) | 87.8 (73.8-95.9) |
| Other reason | N | 54 | 46.3 | 64.0 (42.5-82.0) | 100 (88.1-100) | 100 (79.4-100) | 76.3 (59.8-88.6) |
| | OP-N | 15 | 60.0 | 55.6 (21.2-86.3) | 100 (54.1-100) | 100 (47.8-100) | 60.0 (26.2-87.8) |
| **Clinitest** | | | | | | | |
| Reason for testing: | | | | | | | |
| Positive self-test | N | 257 | 95.3 | 85.7 (80.7-89.8) | 91.7 (61.5-99.8) | 99.5 (97.4-100) | 23.9 (12.6-38.8) |
| | OP-N | 290 | 96.6 | 86.1 (81.5-89.9) | 80.0 (44.4-97.5) | 99.2 (97.1-99.9) | 17.0 (7.6-30.8) |
| Symptoms | N | 459 | 54.5 | 69.6 (63.5-75.2) | 99.0 (96.6-99.9) | 98.9 (96.0-99.9) | 73.1 (67.6-78.2) |
| | OP-N | 390 | 57.9 | 76.1 (70.0-81.5) | 98.2 (94.7-99.6) | 98.3 (95.1-99.6) | 74.9 (68.5-80.5) |
| Close contact | N | 144 | 49.3 | 49.3 (37.2-61.4) | 100 (95.1-100) | 100 (90.0-100) | 67.0 (57.3-75.7) |
| | OP-N | 99 | 44.4 | 65.9 (50.1-79.5) | 92.7 (82.4-98.0) | 87.9 (71.8-96.6) | 77.3 (65.3-86.7) |
| Other reason | N | 29 | 34.5 | 70.0 (34.8-93.3) | 100 (82.4-100) | 100 (59.0-100) | 86.4 (65.1-97.1) |
| | OP-N | 24 | 50.0 | 58.3 (29.1-70.9) | 91.7 (61.5-99.8) | 87.5 (47.3-99.7) | 68.8 (41.3-89.0) |

Flowflex=Acon Labs Flowflex COVID-19 Antigen Home Test; MPBio=MP Biomedicals Rapid SARS-CoV-2 Antigen Test Card; Clinitest=Siemens-Healthineers CLINITEST Rapid COVID-19 Antigen Test; NPV=negative predictive value; PPV=positive predictive value; N = nasal sampling; OP-N = combined oropharyngeal and nasal sampling.

**Table S8 User experience of the three Ag-RDTs.** Up to 3% of the participants in the three nasal sampling groups and up to 5.5% in the OP-N sampling groups reported problems with conducting and/or interpreting the self-test. The higher percentage in the OP-N sampling group was primarily explained by fear or uncertainty of placing the swab (far) up the throat. Participants that performed nasal sampling more often indicated they performed and interpreted the test correctly than those that performed OP-N sampling.

| | Flowflex | MPBio | | Clinitest | |
|---|---|---|---|---|---|
| | Nasal sampling | Nasal sampling | Oropharyngeal-nasal sampling | Nasal sampling | Oropharyngeal-nasal sampling |
| | N = 1938 | N = 1706 | N = 543 | N = 1579 | N = 653 |
| Did you experience problems when conducting the test? No. (%) | | | | | |
| Yes | 30 (1.5) | 49 (2.9) | 25 (2.6) | 36 (2.3) | 36 (5.5) |
| I had a problem when performing the test, No. (%) | 23 (76.7) | 36 (73.5) | 21 (84.0) | 30 (83.3) | 29 (80.6) |
| Unsure how deep to place the swab up my nose | 8 (34.8) | 3 (8.3) | 1 (4.8) | 1 (3.3) | 1 (3.4) |
| Unsure how deep to place the swab up my throat | N/A | N/A | 5 (23.8) | N/A | 5 (17.2) |
| Fear of placing the swab too far up my nose | 7 (30.4) | 3 (8.3) | 2 (9.5) | 3 (10.0) | 2 (6.9) |
| Fear of placing the swab too far up my throat | N/A | N/A | 4 (19.0) | N/A | 9 (31.0) |
| Spilled fluid | 1 (4.3) | 1 (2.8) | 0 (0) | 1 (3.3) | 0 (0) |
| Other, unspecified | 13 (56.6) | 33 (91.7) | 8 (38.1) | 27 (90) | 7 (24.1) |
| I had a problem interpreting the test, No. (%) | 3 (10.0) | 6 (12.2) | 2 (8.0) | 5 (13.9) | 4 (11.1) |
| I saw no control line | 1 (33.3) | 6 (100) | 2 (100) | 5 (100) | 4 (100) |
| Liquid didn't travel through the cassette | 1 (33.3) | 2 (33.3) | 1 (50.0) | 5 (100) | 4 (100) |
| Unclear whether the test was positive | 3 (100) | 2 (33.3) | 1 (50.0) | 1 (20.0) | 2 (50.0) |
| Other, unspecified | 1 (33.3) | 2 (33.3) | 2 (100) | 4 (80) | 2 (50.0) |
| The user manual was clear, No. (%) | | | | | |
| Agree | 1879 (97.0) | 1659 (97.2) | 516 (95.0) | 1323 (83.8) | 575 (88.1) |
| Neutral | 42 (2.2) | 34 (2.0) | 20 (3.7) | 110 (7.0) | 38 (5.8) |
| Disagree | 11 (0.6) | 8 (0.5) | 5 (0.9) | 144 (9.1) | 37 (5.7) |
| Unknown | 2 (0.1) | 5 (0.3) | 2 (0.4) | 2 (0.1) | 3 (0.5) |
| I am sure I performed the test correctly, No. (%) | | | | | |
| Agree | 1838 (94.8) | 1598 (93.7) | 490 (90.2) | 1525 (96.6) | 584 (89.4) |
| In doubt | 90 (4.6) | 97 (5.7) | 48 (8.8) | 50 (3.2) | 60 (9.2) |
| Disagree | 4 (0.2) | 5 (0.3) | 3 (0.6) | 2 (0.1) | 5 (0.8) |
| Unknown | 6 (0.3) | 6 (0.3) | 2 (0.4) | 2 (0.1) | 4 (0.6) |
| I am sure I interpreted the test correctly, No. (%) | | | | | |
| Agree | 1915 (98.8) | 1671 (98.0) | 525 (96.7) | 1555 (98.5) | 634 (97.1) |

| | | | | | |
|---|---|---|---|---|---|
| In doubt | 12 (0.6) | 23 (1.3) | 13 (2.4) | 19 (1.2) | 14 (2.1) |
| Disagree | 3 (0.2) | 5 (0.3) | 3 (0.6) | 3 (0.2) | 1 (0.2) |
| Unknown | 8 (0.4) | 7 (0.4) | 2 (0.4) | 2 (0.1) | 4 (0.6) |

*N/A = not applicable*

**Table S9 Follow-up information of participants that were tested again within 10 days after the initial test.** Follow-up information was not available for 56/211 (26.5%), and 110/425 (25.9%), and 98/297 (33.0%) of nasal sampling participants with Flowflex, MPBio, and Clinitest and 45/179 (25.1%) and 79/231 (34.2%) of OP-N sampling participants with MPBio and Clinitest, that had an Ag-RDT result available and were tested negative with RT-PCR at their initial test site visit, respectively. Of the remaining participants 75/155 (48.4%), 161/315 (51.1%), and 100/199 (50.3%), and 61/134 (45.5%) and 68/152 (44.7%) were not tested within 10 days after the initial test, respectively. Overall, follow-up information was available for 71.1% (955/1343) of participants with a negative RT-PCR test result at baseline. Of this group, 51.3% (490/955) reported to have been re-tested within 10 days, with 16.3% of them (80/490) testing positive.

| Test result available from the reference test and… | Flowflex | MPBio | | Clinitest | |
|---|---|---|---|---|---|
| Type of sampling | Nasal | Nasal | OP-N | Nasal | OP-N |
| Sample size (Test during 10-day follow-up) | N = 80/155 (51.6%) | N = 154/315 (48.9%) | N = 73/134 (54.5%) | N = 99/199 (49.7%) | N = 84/152 (55.3%) |
| Type of test, No. (%) | | | | | |
| public health service COVID-19 test sites | 25 (31.3) | 50 (32.5) | 26 (35.6) | 36 (29.5) | 29 (34.5) |
| RT-PCR test | 23 (92.0) | 45 (90.0) | 24 (92.3) | 33 (91.7) | 25 (86.2) |
| Ag-RDT | 0 (0) | 2 (4.0) | 1 (3.8) | 0 (0) | 1 (3.4) |
| Unknown | 2 (8.0) | 3 (6.0) | 1 (3.8) | 3 (8.3) | 3 (10.3) |
| Other test site (incl. at work) | 5 (6.3) | 6 (3.9) | 3 (4.1) | 3 (2.5) | 4 (4.8) |
| RT-PCR test | 2 (40.0) | 4 (66.7) | 1 (33.3) | 1 (33.3) | 2 (50.0) |
| Ag-RDT | 3 (60.0) | 2 (33.3) | 2 (66.7) | 2 (66.7) | 2 (50.0) |
| Unknown | 0 (0) | 0 (0) | 0 (0) | 0 (0) | 0 (0) |
| Hospital or general practitioner test site | 2 (2.5) | 1 (0.6) | 0 (0) | 1 (0.8) | 0 (0) |
| RT-PCR test | 1 (50.0) | 0 (0) | N/A | 0 (0) | N/A |
| Ag-RDT | 1 (50.0) | 0 (0) | N/A | 1 (100) | N/A |
| Unknown | 0 (0) | 1 (100) | N/A | 0 (0) | N/A |
| Self-test | 68 (85.0) | 119 (77.3) | 61 (83.6) | 80 (80.8) | 74 (88.1) |
| Positive test result, No. (%) | 11 (13.8) | 19 (12.3) | 16 (22.2) | 18 (18.4) | 16 (19.0) |

OP-N = oropharyngeal and nasal; N/A = not applicable
