## Supplementary figures and images for "Accuracy of COVID-19 self-tests with unsupervised nasal or nasal plus oropharyngeal self-sampling in symptomatic individuals in the Omicron period"

### Supplementary Figure 1

Flowflex (*Rotterdam*)

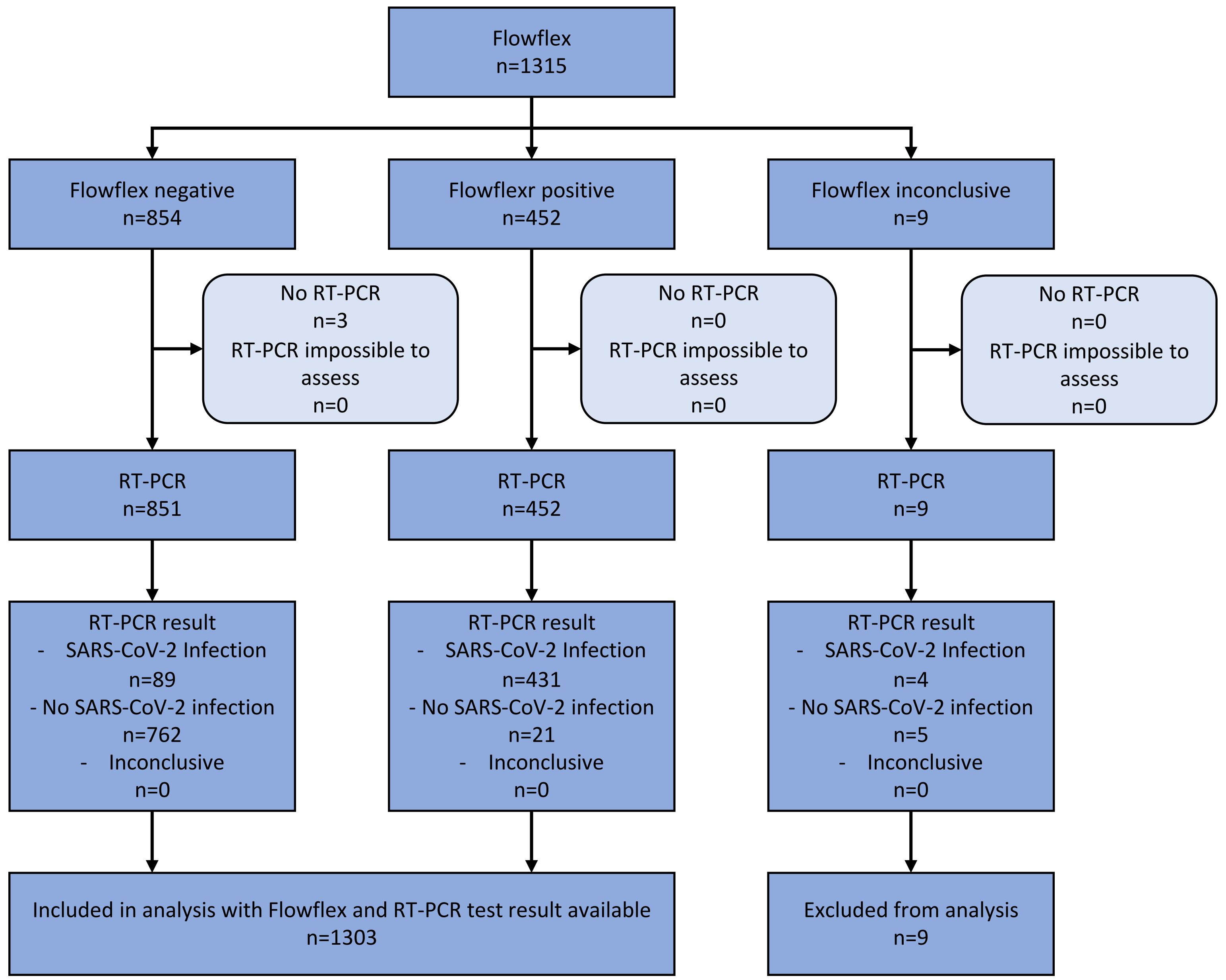


MPBio (*Tilburg*)

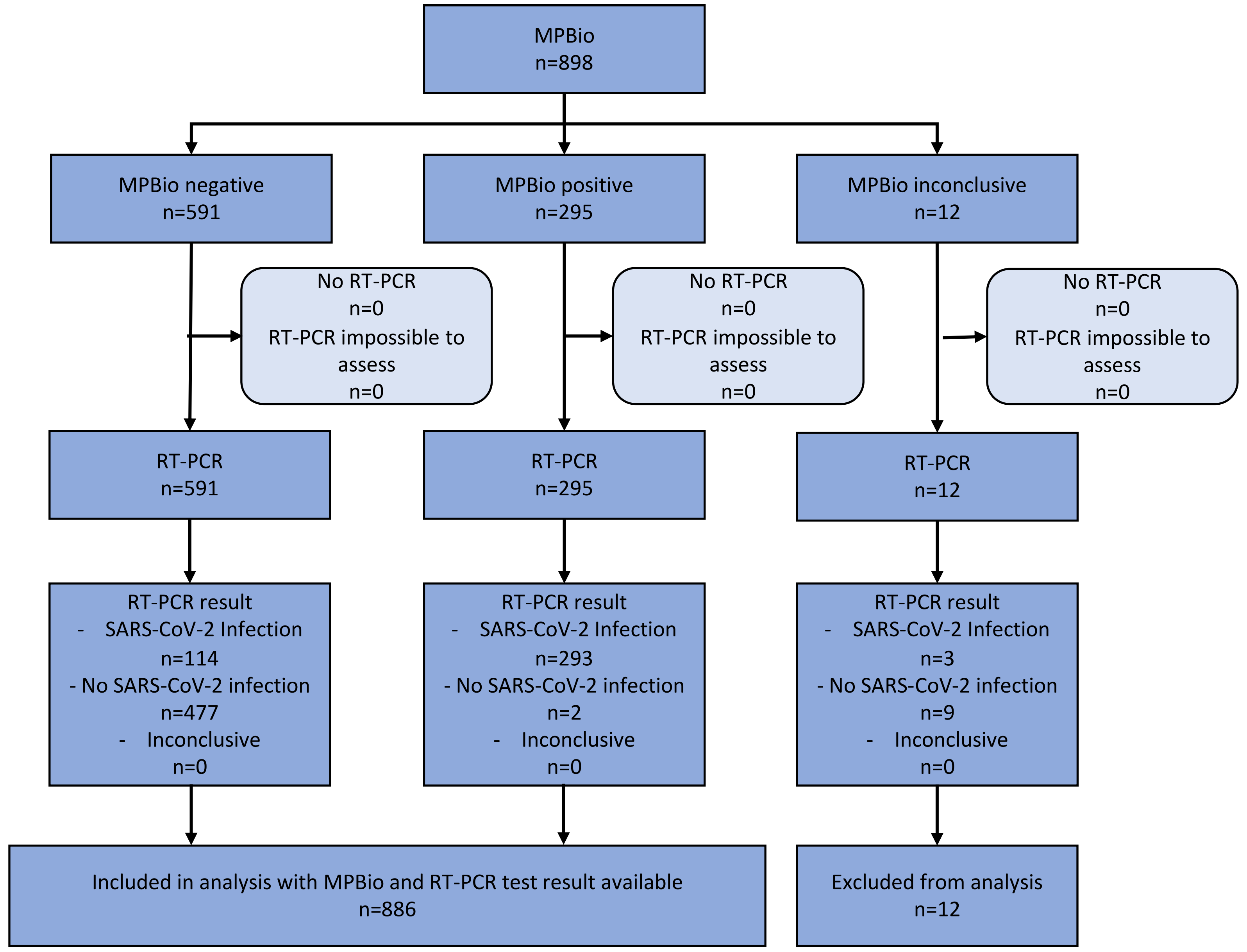


Clinitest (*Roosendaal*)

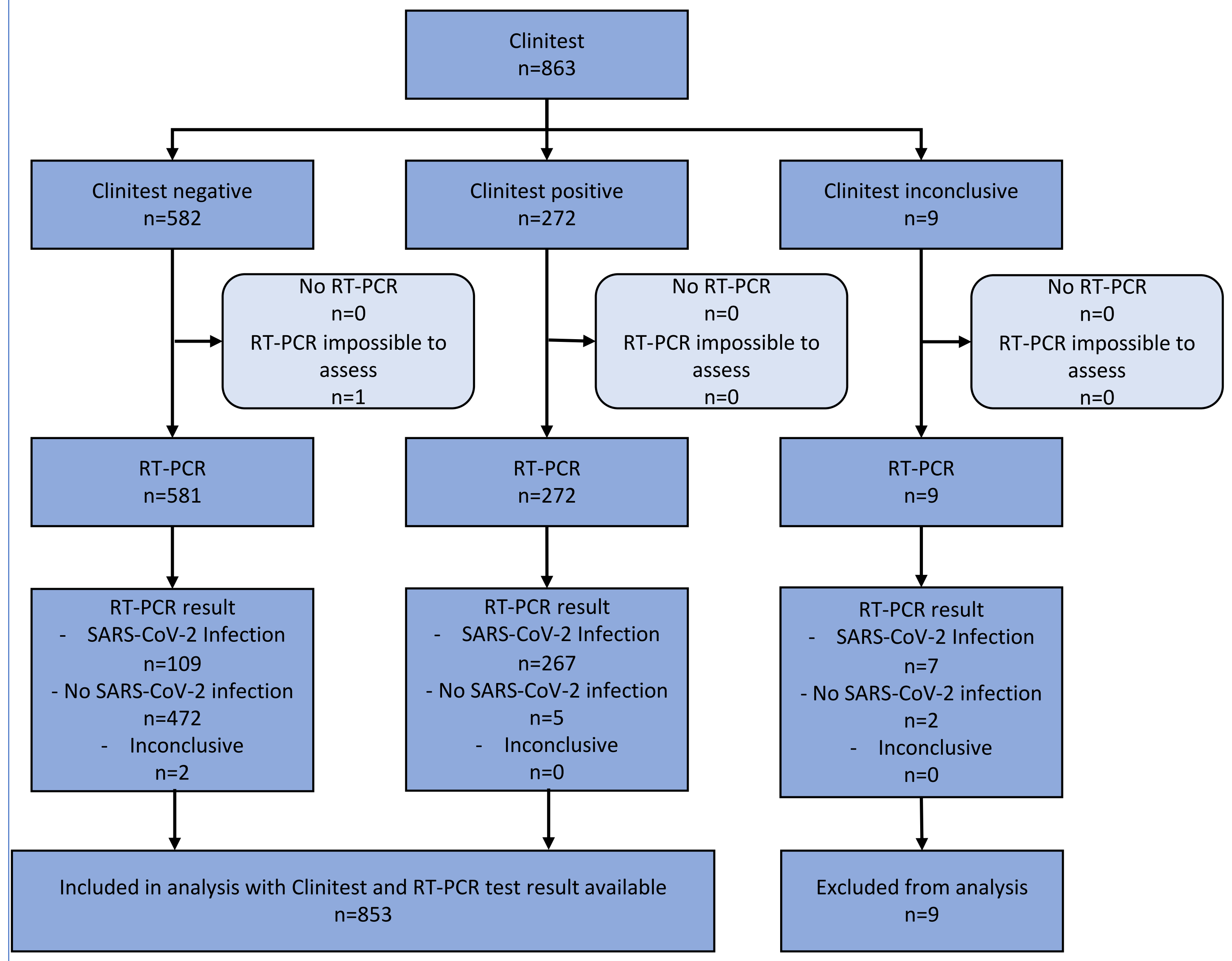

### Supplementary Figure 2

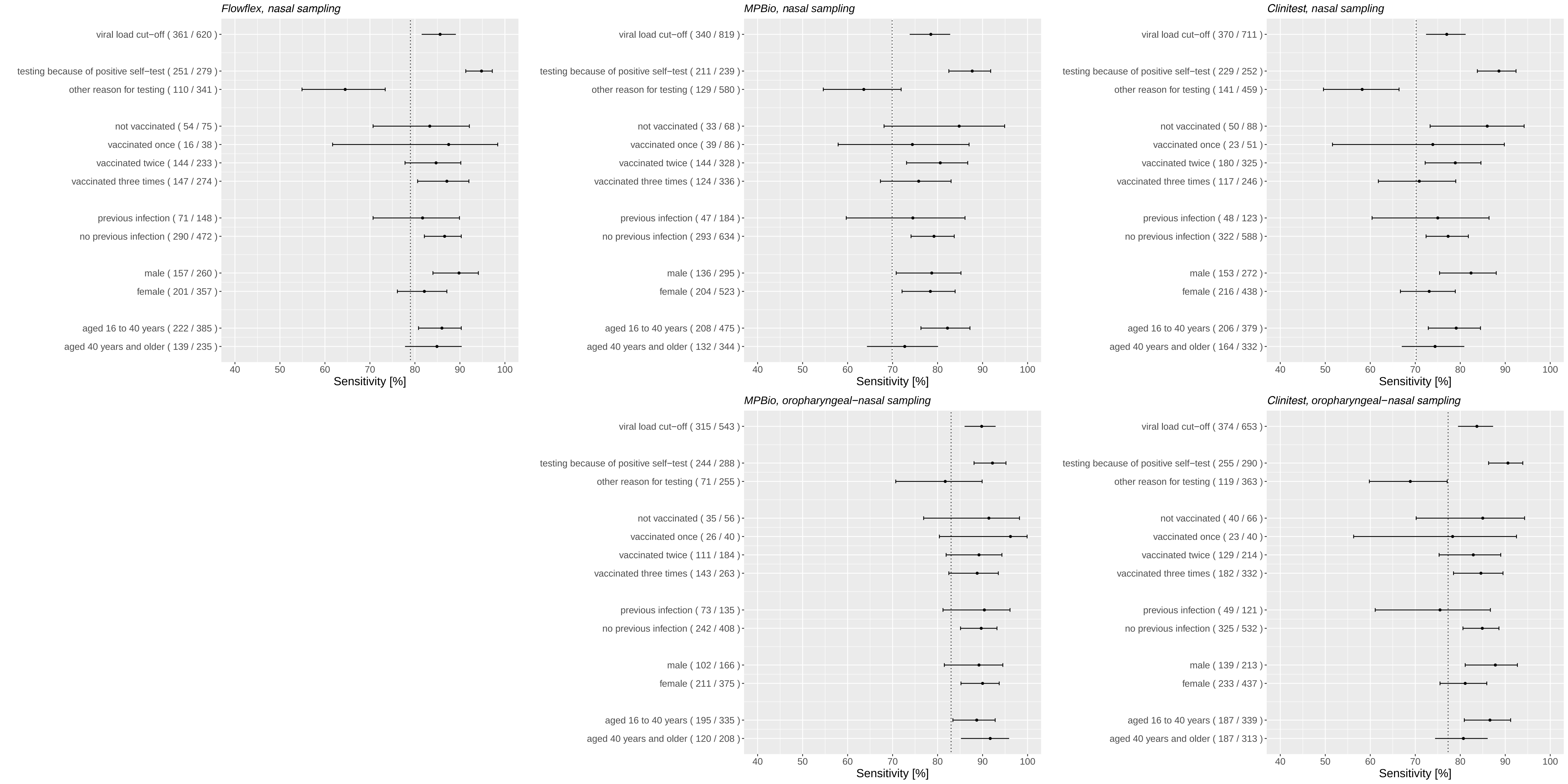

### Supplementary Figure 3

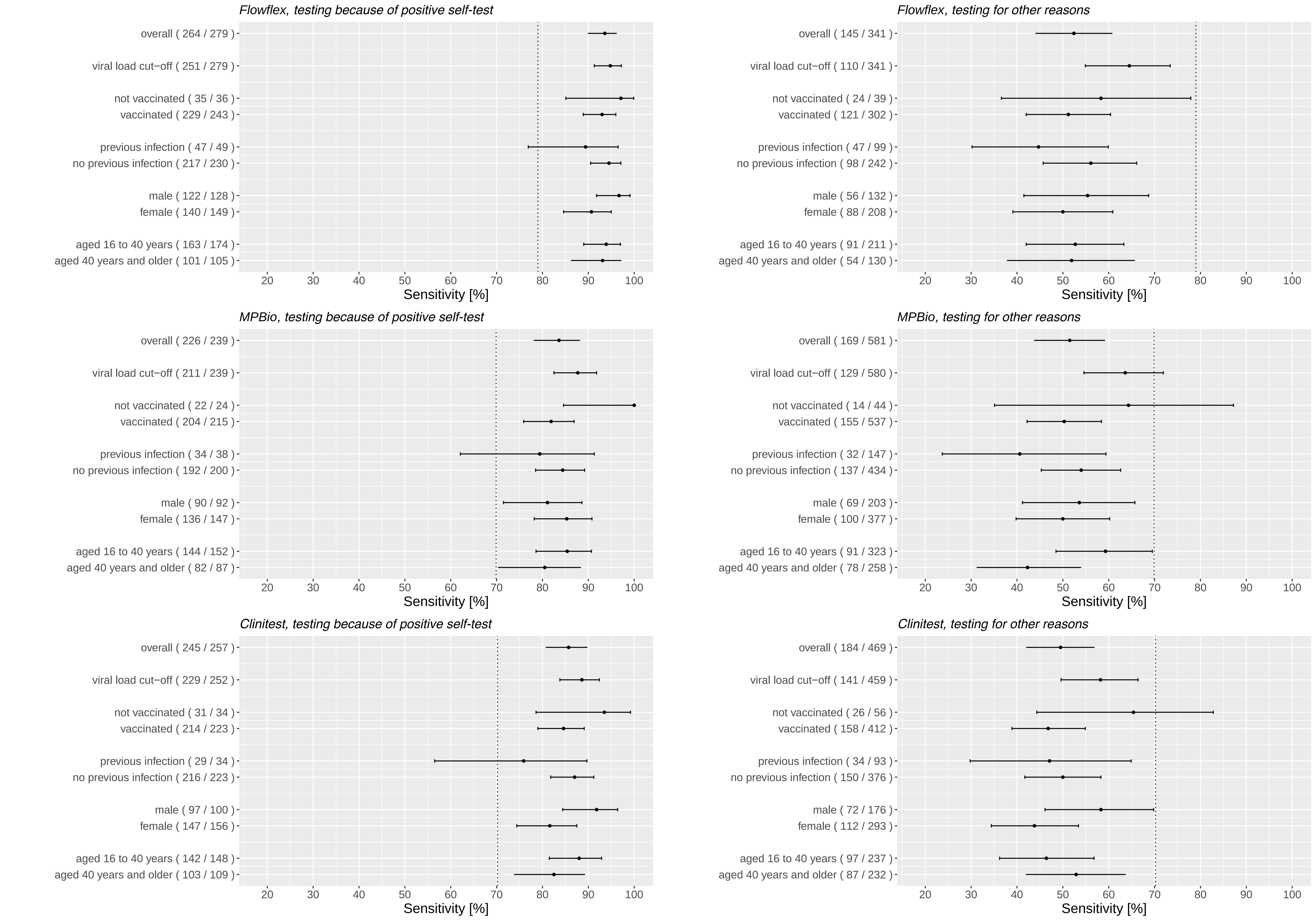

### Supplementary Figure 4

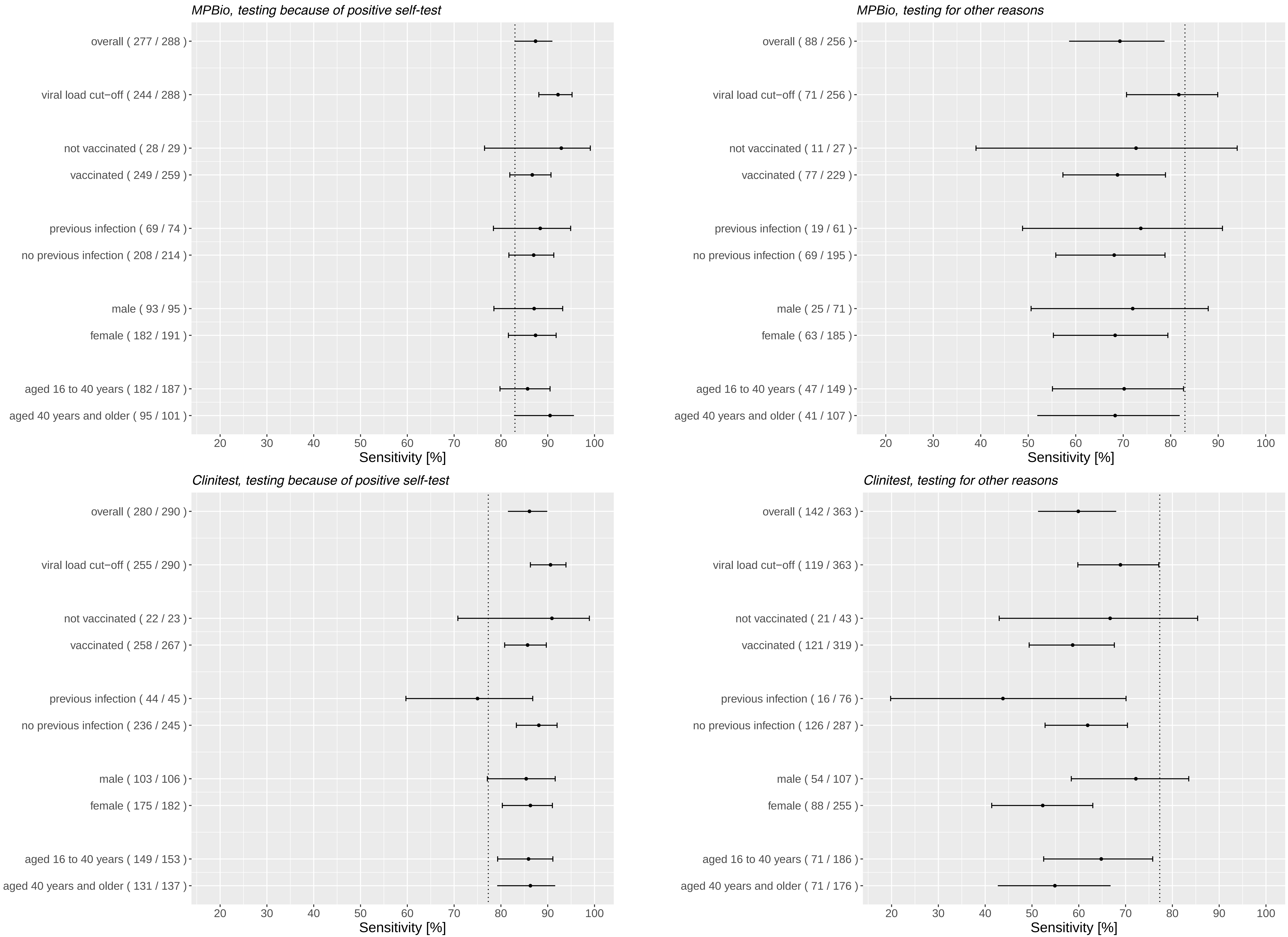

### Supplementary Figure 5

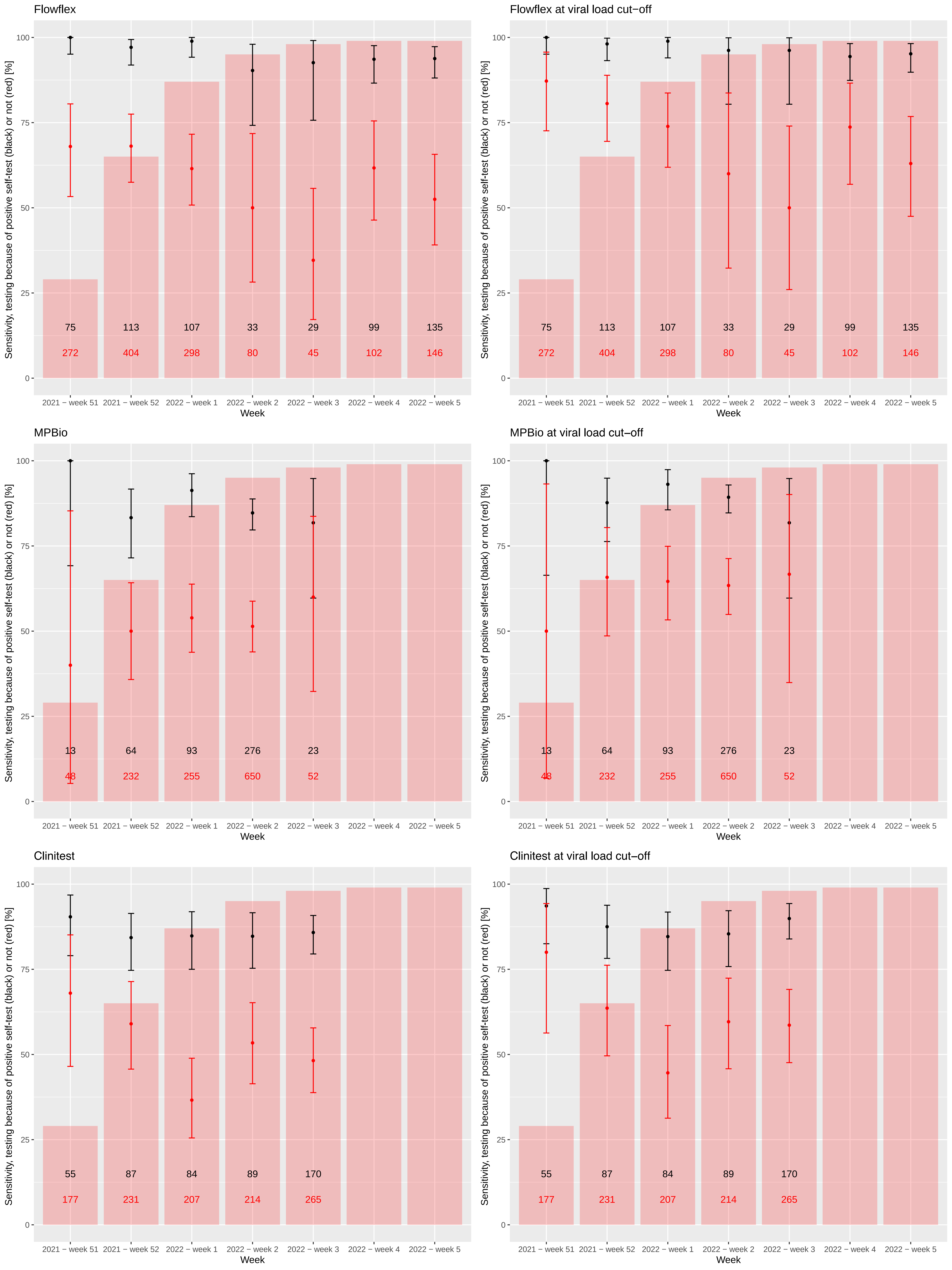
